# Risks of digestive diseases in long COVID: Evidence from a large-scale cohort study

**DOI:** 10.1101/2023.04.25.23289080

**Authors:** Yuying Ma, Lijun Zhang, Rui Wei, Weiyu Dai, Ruijie Zeng, Dongling Luo, Rui Jiang, Huihuan Wu, Zewei Zhuo, Qi Yang, Jingwei Li, Felix W Leung, Chongyang Duan, Weihong Sha, Hao Chen

**Author notes:** ***Correspondence:*** Prof. Felix W Leung, David Geffen School of Medicine, University of California Los Angeles, Los Angeles 90024, California, USA;, Prof. Chongyang Duan, Department of Biostatistics, School of Public Health, Southern Medical University, Guangzhou 510000, China;, Prof. Weihong Sha, Department of Gastroenterology, Guangdong Provincial People’s Hospital (Guangdong Academy of Medical Sciences), Southern Medical University, Guangzhou 510080, China;, Prof. Hao Chen, Department of Gastroenterology, Guangdong Provincial People’s Hospital (Guangdong Academy of Medical Sciences), Southern Medical University, Guangzhou 510080, China. These authors contributed equally to this work (Yuying Ma, Lijun Zhang, Rui Wei, Weiyu Dai).

## Abstract

**Objectives:** This study aims to evaluate the effect of coronavirus disease 2019 (COVID-19) on the long-term risk of digestive diseases in the general population.

**Design:** Large-scale population-based cohort study based on a prospective cohort.

**Setting:** UK Biobank cohort linked to multiple nationwide electronic health records databases.

**Participants:** The cohort consisted of 112,311 individuals who survived the initial 30 days following severe acute respiratory syndrome coronavirus 2 (SARS-CoV-2) infection, as well as two control groups: a contemporary group (n = 359,671) without any history of COVID-19, and a historical control group (n = 370,979) that predated the COVID-19 outbreak.

**Main outcome measures:** Main outcomes were predefined digestive diseases. Hazard ratios and corresponding 95% confidence intervals (CI) were computed utilizing the Cox regression models after inverse probability weighting.

**Results:** Compared with the contemporary control group, patients with previous COVID-19 infection had higher risks of digestive diseases, including functional gastrointestinal disorders (hazard ratios [HR] 1.95 (95% CI 1.62 to 2.35)); peptic ulcer disease (HR 1.27 (1.04 to 1.56)); gastroesophageal reflux disease (GERD) (HR 1.46 (1.34 to 1.58)); inflammatory bowel diseases (HR 1.40 (1.02 to 1.90)); gallbladder disease (HR 1.28 (1.13 to 1.46)); severe liver disease (HR 1.46 (1.12 to 1.90)); non-alcoholic liver disease (HR 1.33 (1.15 to 1.55)); and pancreatic disease (HR 1.43 (1.17 to 1.74)). The risks of GERD were stepwise increased with severity of the acute phase of COVID-19 infection. The results were consistent when using the historical cohort as the control group.

**Conclusions:** Our study provides important insights into the association between COVID-19 and the long-term risk of digestive system disorders. COVID-19 patients are at a higher risk of developing gastrointestinal disorders, with stepwise increased risk with the severity and persisting even after one year follow-up.

## 1. Introduction

The ongoing pandemic caused by the severe acute respiratory syndrome coronavirus 2 (SARS-CoV-2), commonly referred to as coronavirus disease 2019 (COVID-19), has become a serious global public health concern ^1^. As we move into the post-pandemic era, there is growing global attention towards the enduring consequences of COVID-19.

COVID-19 mainly involves the respiratory system, together with multisystem. During the recovery period, many patients suffer prolonged symptoms, such as dyspepsia, chest pain, fatigue, cognitive impairment, anxiety, and depression ^2^. The Centers for Disease Control and Prevention defines residual symptoms after four weeks of infection as long COVID or post-COVID conditions ^3^. COVID patients who survived the acute phase of COVID-19 are at an increased risk of developing cardiovascular diseases ^4^, mental health disorders^5^, kidney dysfunctions^6^ and metabolic diseases in the long term^7 8^. Therefore, it is crucial to pay more attention to post-acute COVID-19 syndrome and its associated complications.

Many COVID-19 patients exhibit gastrointestinal symptoms, with diarrhea, abdominal pain, nausea, vomiting, and anorexia being the most common ones^9^. The mechanisms underlying these manifestations of COVID-19 are complex and multi-factorial, involving direct viral invasion, immune-mediated tissue damage, dysregulation of angiotensin-converting enzyme 2 (ACE2), and gut dysbiosis^10^. However, there is still a lack of evidence on the long-term risks of digestive outcomes in COVID-19 patients during the post-recovery phase. Several studies have demonstrated that the risks of digestive diseases were increased in COVID-19 patients (ref). But existing studies were mainly limited to hospitalized patients of severe and critical COVID, small sample sizes, short follow-up duration, single-center design, lack of controls or sex-selective bias, which greatly affected the generalizability and accuracy of their results^11-14^. Additionally, vaccination against COVID-19 may help reduce the risk of complications from the disease. This issue has not been well resolved in previous studies^15^. In this study, we designed a large-scale population-based, multi-center cohort study based on a prospective cohort from UK Biobank to investigate the long-term hazard of digestive diseases among COVID patients. We also estimated the risk according to different follow-up times, the severity of the acute COVID-19 infection to make up for those limitations of previous studies.

## 2. Methods

### 2.1 Study population and design

The UK Biobank database recruited 502 368 participants aged 37 to 73 years from the general population between 2006 and 2010 throughout the United Kingdom. It provided information on health-related aspects through extensive baseline questionnaires, verbal interviews, and physical measurements. Hospital inpatient data were updated regularly by linking to Hospital Episode Statistics (HES) for England, Scottish Morbidity Record for Scotland, and Patient Episode Database for Wales. Death data are obtained through linkage to National Health Service (NHS) Digital and NHS Central Register. For the purpose of conducting COVID-19 related research, the UK Biobank has provided records of COVID-19 testing results (using RT-PCR testing on nasopharyngeal swab specimens through linkage to Public Health England (PHE). Updated primary care data from two major General Practice (GP) data system providers, EMIS and TPP, have also been obtained for approximately 450,000 UK Biobank participants in England (from 1938 to October 31th, 2022). Further details about the UK Biobank can be found elsewhere^16^. UK Biobank study obtained written informed consent from each participant and ethical approval from the North West Multi-Centre Research Ethics Committee (approval numbers: 11/NW/0382, 16/NW/0274, and 21/NW/0157). The current study has been approved by the UK Biobank project 83339.

To investigate the impact of COVID-19, two control cohorts - historical and contemporary controls were conducted. The historical controls started from January 31^st^, 2017 and ended on October 30^th^, 2019 (3 years). Participant died before January 30th, 2020, the outbreak of COVID-19 (n = 28,980) and those lost to follow-up (n = 1, 298) were excluded.

To ensure a similar distribution of follow-up time between the contemporary groups and COVID-19 groups, we randomly assigned the start time of follow-up for the contemporary control group based on the start time of follow-up for the COVID-19 group. Meanwhile, the start of follow-up time for the historical control group were also randomly assigned to be three years prior to the time of COVID-19 infection in COVID-19 group.

### 2.2 Definition of COVID-19 infection

COVID-19 infection was defined as the first positive result on COVID-19 polymerase chain reaction (PCR) testing or patients firstly diagnosed with COVID-19 (U07.1 and U07.2). Public Health England (PHE), Public Health Scotland, and Secure Anonymous Information Linkage linked with the UK Biobank to provide COVID-19 testing results from Pillar 1 (swab testing of patients with clinical need or serving as healthcare professionals in PHE laboratories and NHS hospitals) and Pillar 2 (swab testing of the wider population). Moreover, participants who were hospitalized due to COVID-19 but without COVID-19 test reports were also considered as COVID-19 cases and the diagnosis date was defined as the admission time. As using hospitalization records alone as a proxy for severity of infection is not a very reliable or nuanced measure, severe COVID-19 cases were defined as patients with a critical care admission within 7 days of COVID-19 diagnosis and/or receipt of invasive or non-invasive mechanical ventilation or other respiratory support treatments, based on previous studies^17^.

### 2.3 Definition of outcomes

As shown in **Supplementary Table 1**, all the outcomes were defined using the 10th revision of the International Classification of Diseases (ICD-10). The outcomes include (1) functional gastrointestinal disorders (FGID): dyspepsia, irritable bowel syndrome (IBS) and constipation; (2) peptic ulcer disease (PUD): gastric ulcer, duodenal ulcer and peptic ulcer (site unspecified); (3)gastro-oesophageal reflux disease (GERD); (4) inflammatory bowel disease (IBD): Crohn’s disease (CD) and ulcerative colitis (UC); (5) severe liver disease: liver failure, hepatic sclerosis or cirrhosis and complication of liver disease ; (6) non-alcohol fatty liver disease (NAFLD); (7) gallbladder disease: cholelithiasis and cholecystitis; (8) pancreatic disease: acute pancreatitis, chronic pancreatitis, pancreatic cyst and other diseases of pancreas.

Cases were defined as the onset of symptoms 30 days or more after the diagnosis of SARS-CoV-2 infection or the index date.

### 2.4 Covariates

In this study, covariates were obtained from baseline data self-reported by participants in verbal interview and periodically updated disease diagnosis data. Pre-defined covariates were selected based on previous knowledge and examination of a Directed Acyclic Graph (DAG)^18-20^(**Supplementary Figure 1**). The covariates included sociodemographic characteristics (age, sex, ethnicity, household income, Townsend Deprivation Index), lifestyle factors (smoking status, alcohol consumption, moderate physical activity) and comorbidities (hypertension, diabetes, heart failure, renal failure, myocardial infarction, asthma, dementia, stroke, chronic obstructive pulmonary disease [COPD]) and history of previous digestive diseases. We calculated the number and proportion of missing data for the covariates and performed multiple imputation using chained equations (MICE packages in R) with 50 iterations to handle the missing data^21^.

### 2.5 Statistical analyses

Baseline characteristics of the COVID-19, historical and contemporary control group were presented as mean (standard deviation) and numbers (percentages) as appropriate. Standardized mean differences between groups were also described.

Inverse probability weights (IPTW) were calculated for each participant to eliminate the impact of confounding factors. To test the effectiveness of weighting, we evaluated the standardized mean differences (SMD) of covariates between the weighted population. A SMD of less than 0.1 is considered to indicate adequate balance in covariates between groups. Cox regression models were then constructed using the inverse probability weights and additionally adjusted for those unbalanced covariates to evaluate the long-term impact of COVID-19 infection on the study outcome.

The weighted cumulative hazards plots were used to visualize the proportional hazards assumption of the Cox models constructed of the contemporary control group and the COVID-19 group. The comparison between the COVID-19 group and historical control group is additionally examined as one of the sensitivity analyses.

In addition, to further investigate the long-term effects of COVID-19 infection, sensitivity analyses were conducted by redefining the outcome as digestive diseases occurring within six months, between six months and one year, and after one year of follow-up. To evaluate whether there was any dose-dependent association of COVID-19 severity with digestive outcomes, we grouped the patients into non-hospitalized COVID, hospitalized COVID and severe COVID-19 according to the COVID-19 severity.

Since complete data on vaccine status were unavailable, sensitive analysis were constructed by restricting the inclusion period of COVID-19 cohort to the period when vaccines were still not available in the UK before December 2020. Hence, the COVID-19 cohort was comprised of people diagnosed with COVID-19 until 30 November 2020 to avoid the effect of vaccination on long-term risk of COVID-19.

Subgroup analyses consisting of sex (male, female), age (< 60 years or ≥ 60 years), race (White or other), obesity (BMI < 30 kg/m^2^ or BMI ≥ 30kg/m^2^), smoking status, alcohol consumption, hypertension, diabetes, heart failure, renal failure, myocardial infarction, COPD, asthma, dementia, stroke were conducted using the same methods as the main analysis to reduce the impact of confounding factors. We selected subgroups based on sociodemographic characteristics, lifestyle, and special disease status.

All analyses were performed using RStudio and R 4.2.1 software. Two-tailed P values less than 0.05 were considered statistically significant.

## 3. Results

### 3.1 Baseline characteristics

**Supplementary Figure 2** demonstrates the criteria used for selecting study cohorts. The COVID-19 group consisted of 112,311 participants, the contemporary control group had 359,671 participants, and the historical control group had 370,979 participants. The median follow-up time was 254 (interquartile range[IQR] 184 – 366) days for the COVID-19 group, 254 (184 – 366) days for the contemporary control group, and 254 (IQR 184 – 367) days for the historical control group. The number and percentage of missing covariates are shown in **Supplementary Table 2**.

### 3.2 Risks of digestive diseases in long COVID

#### COVID-19 group versus contemporary control group

The baseline characteristics of the study population before and after weighting are presented in **Table 1** and **Supplementary Table 3**. After weighting, all characteristics were well balanced between the groups.

**Table 1.** Baseline characteristics of COVID-19 group and contemporary controls after weighting

| Characteristics | COVID-19 group<br>(n = 112 311) | Contemporary controls<br>(n = 359 671) | SMD |
| --- | --- | --- | --- |
| Age, mean(SD), years | 56.2(8.1) | 56.2(8.1) | <0.001 |
| Sex, female, n(%) | 61 546(54.8) | 198 538(55.2) | 0.008 |
| Ethnicity, White, n(%) | 10 6134(94.5) | 339 889(94.5) | 0.001 |
| Household income |  |  | 0.003 |
| <18 000, n(%) | 24 371(21.7) | 78 049(21.7) |  |
| 18 000-30 999, n(%) | 28 527(25.4) | 91 716(25.5) |  |
| 31 000-51 999, n(%) | 29 650(26.4) | 94 953(26.4) |  |
| 52 000-100 000, n(%) | 23 361(20.8) | 74 452(20.7) |  |
| >100 000, n(%) | 6 514(5.8) | 20 501(5.7) |  |
| Deprivation index, mean(SD) | -1.3(3.0) | -1.3(3.1) | 0.001 |
| BMI, mean(SD), kg/m<sup>2</sup> | 27.4(4.7) | 27.4(4.8) | 0.001 |
| Alcohol consumption |  |  | 0.005 |
| Daily or almost daily, n(%) | 22 799(20.3) | 72 654(20.2) |  |
| Three or four times a week, n(%) | 26 168(23.3) | 83 803(23.3) |  |
| Once or twice a week, n(%) | 29 089(25.9) | 93 514(26) |  |
| One to three times a month, n(%) | 12 579(11.2) | 40 283(11.2) |  |
| Special occasions only or never, n(%) | 12 803(11.4) | 41 002(11.4) |  |
| Never, n(%) | 8 873(7.9) | 28 414(7.9) |  |
| Smoking status |  |  | 0.002 |
| Never smoker, n(%) | 62 557(55.7) | 200 696(55.8) |  |
| Previous smoker, n(%) | 38 523(34.3) | 123 007(34.2) |  |
| Current smoker, n(%) | 11 343(10.1) | 35 967(10) |  |
| Physical activity, mean(SD), MET minutes/week | 2645.2(2702.9) | 2650.1(2708.0) | 0.002 |
| Comorbidities |  |  |  |
| Hypertension, n(%) | 40 657(36.2) | 129 841(36.1) | 0.002 |
| Diabetes, n(%) | 8 648(7.7) | 27 335(7.6) | 0.002 |
| Renal failure, n(%) | 4 717(4.2) | 15 106(4.2) | 0.002 |
| Myocardial infarction, n(%) | 4 829(4.3) | 15 466(4.3) | 0.002 |
| Stroke, n(%) | 3 032(2.7) | 9 351(2.6) | 0.003 |
| COPD, n(%) | 5 054(4.5) | 14 387(4) | 0.025 |
| Asthma, n(%) | 15 611(13.9) | 49 635(13.8) | 0.002 |
| Heart failure, n(%) | 2 583(2.3) | 7 553(2.1) | 0.013 |
| Dementia, n(%) | 786(0.7) | 2 518(0.7) | 0.002 |
| History of previous digestive diseases, n(%) | 36 950(32.9) | 117 612(32.7) | 0.003 |

SMD: standard mean difference; BMI: body mass index; MET: metabolic equivalent of task;
COPD: chronic obstructive pulmonary disease; SD: standard deviation

Figure 1. provides the risks of digestive outcomes in these groups. Compared to the contemporary control group, people who survived the first 30 days of COVID-19 showed an increased risk of FGID (HR 1.95 (95% confidence interval [CI] 1.62 to 2.35), P < 0.001); PUD (HR 1.27 (1.04 to 1.56), P = 0.021); GERD (HR 1.46 (1.34 to 1.58), P < 0.001); IBD (HR 1.40 (1.02 to 1.90), P = 0.035); gallbladder disease (HR 1.28 (1.13 to 1.46), P < 0.001); severe liver disease (HR 1.46 (1.12 to 1.90), P = 0.005); NAFLD (HR 1.33 (1.15 to 1.55), P < 0.001); and Pancreatic disease (HR 1.43 (1.17 to 1.74), P < 0.001). The weighted cumulative hazards plots are shown in **Figure 2**.

**Figure 1.**
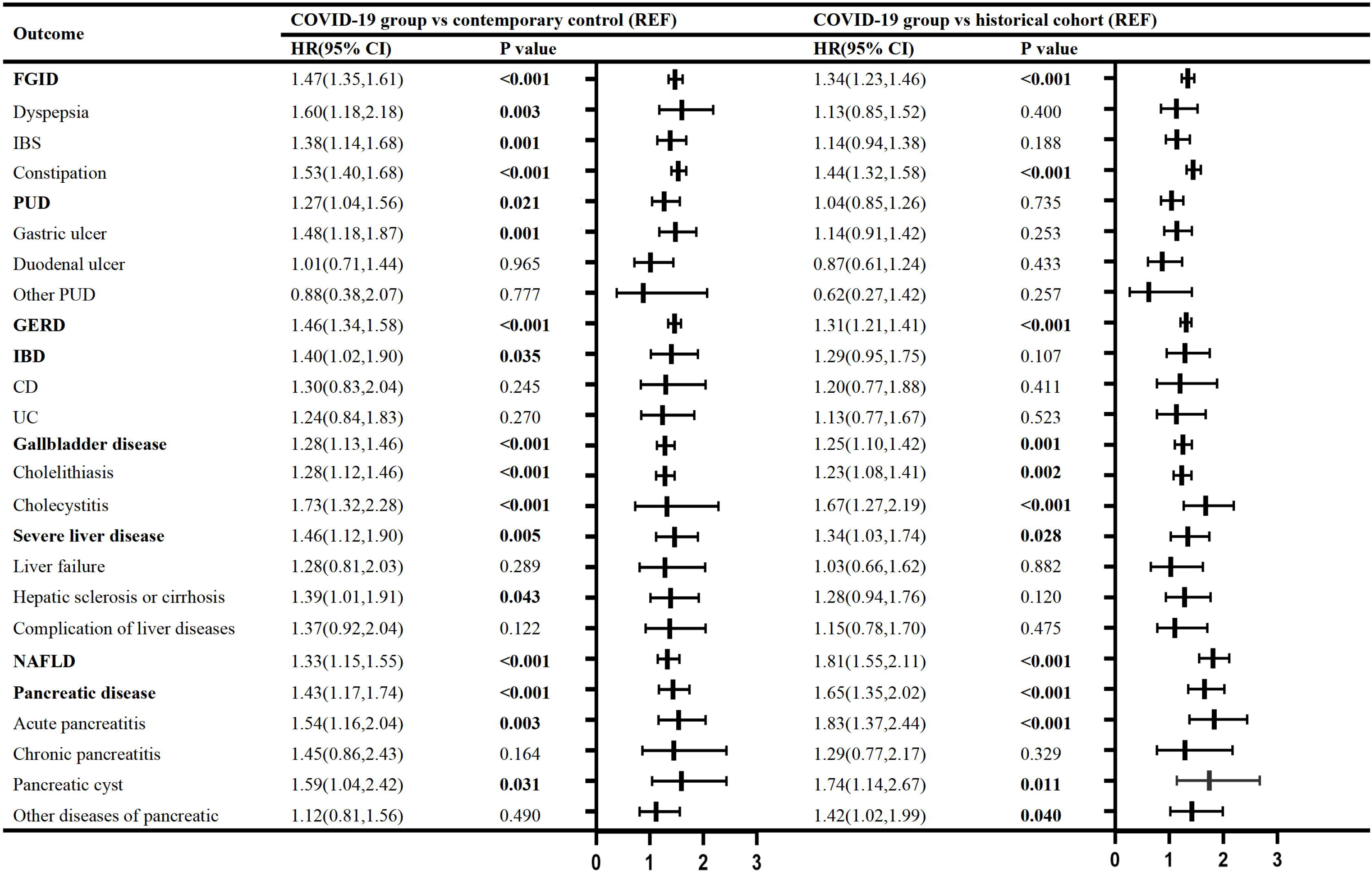
Hazard ratio of digestive outcomes in COVID-19 group compared to the contemporary and historical controls. HR: hazard ratios; CI: confidence interval; Outcomes were ascertained 30 days after the COVID-19-positive test until the end of follow-up. Weighted HRs after IPTW and 95% CIs are presented.

**Figure 2.**
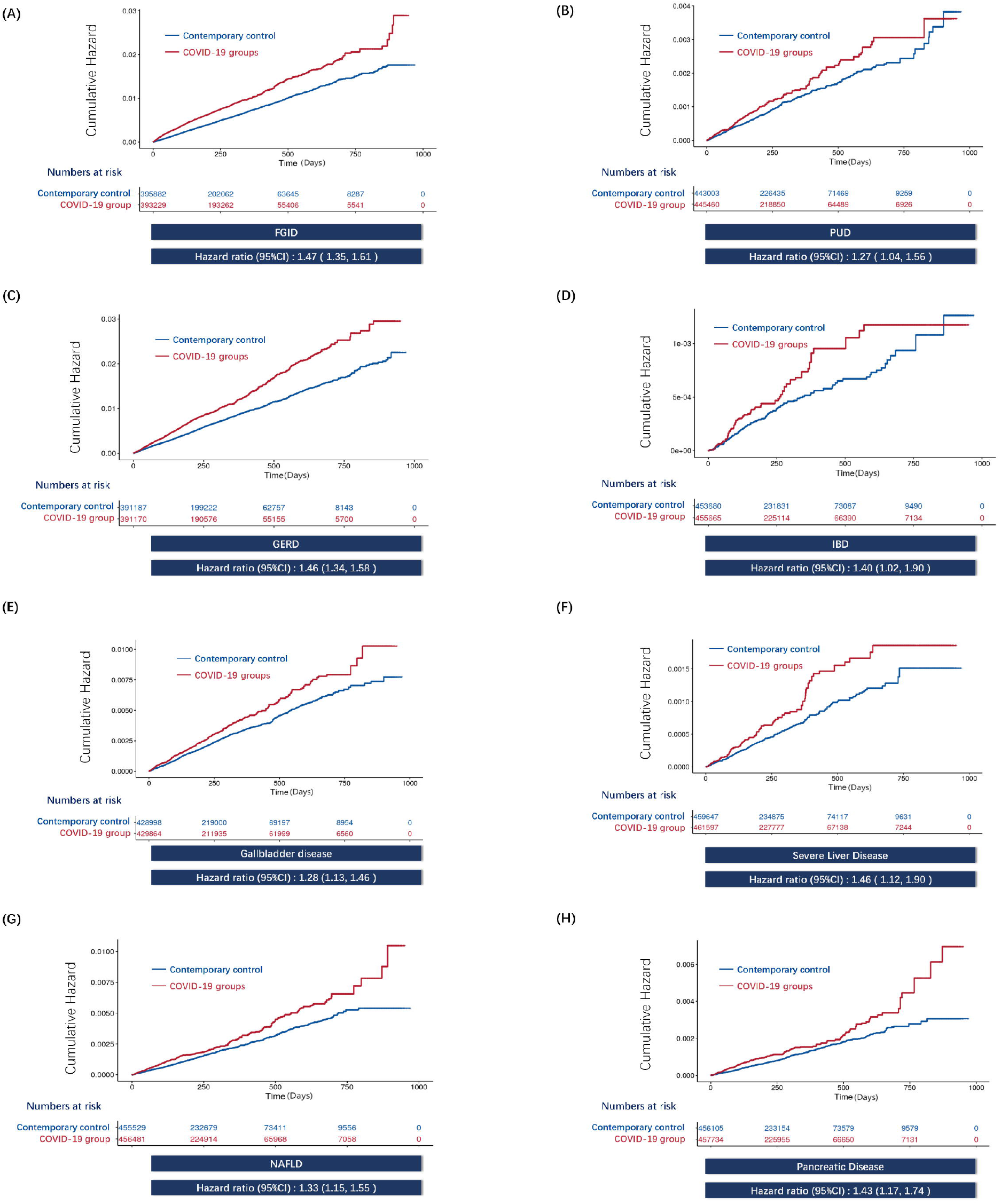
IPTW-weighted cumulative hazards plots of COVID-19 group and contemporary control in digestive outcomes. The cumulative hazard and weighted hazard ratios for FGID **(A)**, PUD **(B)**, GERD **(C)**, IBD **(D)**, gallbladder disease **(E)**, Sever liver diseases **(F)**, NAFLD **(G)** and pancreatic disease **(H)** in COVID-19 group as compared with contemporary control are illustrated. 95% confidence intervals are shown in parentheses.

**Supplementary Table 4** showed the results of COVID-19 group compared with contemporary control group at different follow-up times. We found persistent increased risks of GERD in COVID-19 group even after one year of follow-up. On the contrary, the risks of FGID, PUD, gallbladder disease and pancreatic disease were only observed within 6 months of post-infection.

We further examined the risks of digestive outcomes by the severity of COVID-19 infection during the acute phase (non-hospitalized [n = 104,201], hospitalized [n = 7,523] and severe COVID-19 [n = 588]); Baseline characteristics of these groups before and after weighting are provided in **Supplementary Table 5, 6**. Assessment of covariate balance after application of weights suggested most covariates were well balanced, except for COPD (SMD = 0.165) and heart failure (SMD = 0.112) between hospitalized COVID and contemporary controls; BMI (SMD = 0.193), alcohol consumption (SMD = 0.148), hypertension (SMD = 0.130), COPD (SMD = 0.209) and history of previous digestive diseases (SMD = 0.115) between severe COVID and contemporary controls. Then we adjusted for those covariates that remained unbalanced in the IPTW-weighted Cox models. Compared with the contemporary control group, the risks of FGID and GERD were evident even among those who were not hospitalized during the acute phase of COVID-19 infection (**Supplementary Figure 3**). The risks of GERD also increased in a graded manner in accordance with the severity of the acute infection from non-hospitalized, hospitalized COVID to severe COVID.

In the sensitive analysis, the COVID-19 group consisted of 8,431 participants when restricting to the period before or after vaccination was available in the UK. **Supplementary Table 7, 8** demonstrates baseline characteristics of COVID-19 group and contemporary controls before and after weighting, indicating covariaties were well balanced, except for COPD (SMD = 0.121). The results were basically consistent with the main analyses (**Supplementary Table 9**).

#### COVID-19 group versus contemporary control group in subgroup analyses

In parallel, we examined the risks of digestive outcomes in COVID-19 group compared to contemporary control in pre-specified subgroups. Subgroup analyses found consistent results that the risks of functional gastrointestinal disorders were evident in all subgroups based on age, ethnicity, deprivation index, body mass index, smoking status, alcohol consumption, physical activities, hypertension, diabetes, heart failure, renal failure, myocardial infarction, COPD, asthma, dementia, stroke **(Supplementary Table 10)**.

As shown in **Supplementary Table 11**, no sex-based differences in digestive diseases between COVID-19 patients and control groups was observed in subgroup analyses by sex.

#### COVID-19 group versus historical control group

We tested the reliability of our study design by evaluating the associations between COVID-19 infection and the long-term digestive outcomes in analyses using a historical control group (from an era predating the COVID-19 pandemic) as the reference category. Demographic and health characteristics before weighting and after weighting are available in **Supplementary Table 12-15, 17-18**, suggesting that most covariates were balanced after application of inverse probability weighting. Unbalanced covariates were further adjusted (age (SMD = 0.276) between non-hospitalized COVID and historical controls; age (SMD = 0.207) and COPD (SMD = 0.165) between hospitalized COVID and historical controls; age (SMD = 0.103), sex (SMD = 0.103), BMI (SMD = 0.200), alcohol consumption (SMD = 0.144), hypertension (SMD = 0.135), COPD (SMD = 0.190) and history of previous digestive diseases (SMD = 0.106) between severe COVID and historical controls. The trend of risks was similar with the results of analyses using the contemporary control in comparisons of COVID-19 group (**Figure 1; Supplementary Table 9-11, 16**).

## 4. Discussion

Our study found that COVID-19 was significantly associated with an increased risk of various digestive system diseases. The risks of GERD increased along with the severity of COVID-19 and did not decrease after one-year of follow-up time. This indicates a dose-dependent and long-term effect of COVID-19 and the risks of digestive disorders.

According to our findings, the risks of most digestive outcomes were evident even in people with mild COVID-19 symptoms that did not receive hospitalization treatment. Given that people with mild symptoms takes up over 95% of the COVID-19 population, even a small incidence rate can result in a large number of people being affected. This highlights the importance of not only addressing the individual needs of those with gastrointestinal disorders in the post-COVID era, but also ensuring that healthcare systems are equipped to provide appropriate care to this population.

Results of this study are consistent with the only previous large-scale study based on the US Veteran Health Administration (VHA) database that people with COVID-19 exhibited increased risks of developing FGID, GERD and other digestive diseases. This add evidence to previous findings. However, there exists non-negligible selection bias of US VHA cohort with nearly 90% male of veteran participants. In contrast, this study was conducted in the UK Biobank cohort, including both male and female participants, thus ensuring the generalizabilty of our results.

### Long-term follow-up

In our study, we conducted a follow-up period of up to two years and a half, enabling us to examine the prolonged impacts of COVID-19 on digestive system diseases. We found that the risks of GERD, NAFLD and pancreatic disease did not significantly decrease even after one year of follow-up, implying a persistent effect of COVID-19 on the digestive system. While our study demonstrated only the short-term risks of developing PUD and gallbladder disease in post-COVID. The result is accordant to the study by Rithvik et al. with the conclusion that COVID-19 led to higher number of new onset post infection-FGID at 3 and 6 months of follow-up^14^. Moreover, our study meaningfully extended the finding of previous researches that the risk of developing FGID persisted after one year. This highlights the importance of enhancing our comprehension of the long-term risks of digestive outcomes in individuals with COVID-19 to develop appropriate care strategies during the post-acute phase.

### Potential mechanisms

The mechanisms behind the associations between COVID-19 and digestive diseases are not yet fully understood, but several possibilities have been suggested.

One possibility is the fecal-oral transmission of the virus, leading to viral infection of the digestive tract^22^. Viral-induced gastrointestinal epithelial cell damage can lead to acute digestive symptoms, such as abdominal pain and diarrhea, which is a common symptom in COVID-19 patients. After the acute phase, the virus infection usually triggers irritable bowel syndrome that causes long-term functional gastrointestinal disorders of the digestive tract^23^. In addition, prolonged shedding of viral particles from the gastrointestinal tract is believed to be another contributing factor to some of the gastrointestinal symptoms associated with long COVID syndrome^24^.

Additionally, interactions between the SARS-CoV-2 spike protein and the expression of the ACE2 receptor on digestive tract might also involve in the development of digestive diseases in COVID-19 patients. ACE2 is the major receptor for SARS-CoV-2 spike proteins during infection ^25^. In fact, the epithelium of the gastrointestinal tract expresses higher level of ACE2 than the lung ^25^, which makes it highly susceptible to SARS-CoV-2 infection ^26^. ACE2 also expresses in the biliary tract and pancreas, which may contribute to the increased risk of gallbladder and pancreas diseases after COVID-19^27 28^. Accordingly, it has been discovered that SARS-CoV-2 exists in the bile^29^ and pancreatic cells ^30 31^ of COVID-19 patients. Therefore, we deduce that the strong expression of ACE2 in the gastrointestinal system enables SARS-CoV-2 to invade the gastrointestinal tract, gallbladder, and pancreas directly, which in turn contributes to the relevant consequences observed in patients with COVID-19.

#### Strengths

Our study has several strengths that contribute to the validity and reliability of our findings. First, our study had a long-term follow-up period of up to two and a half years, which allowed us to analyze the sustained effects of COVID-19 on digestive system diseases. This extended follow-up period adds further value to our study, as it provides insight into the long-term implications of COVID-19 on digestive health. Secondly, by using the UK Biobank, a nationwide cohort including both female and male, the results of our study were of high generalizability compared to previous study of US VHA. Thirdly, to enhance the robustness of the results, we employed two control groups including a contemporary and a historical cohort, and considered covariates to minimize selection and confounding bias. Furthermore, we conducted the subgroup analyses restricting to the period before or after vaccination was available to eliminate the impact of vaccination. Fourth, we investigated the dose-response relationship of patients of different COVID-19 severity and digestive diseases.

#### Limitations

Although our study provides important insights into the association between COVID-19 and digestive system diseases, it is not without limitations. Firstly, our study was conducted predominantly on a European population, further studies in other ethnicity are warranted to confirm our results. Secondly, being an observational study, we cannot establish a causal relationship between COVID-19 and the long-term risk of digestive system diseases. However, we observed dose-response relationship between the severity of COVID-19 and risk of several digestive outcomes, which to some extent suggested a possibility for causality. Thirdly, unmeasured confounding bias can not be excluded due to unmeasured confounding factors.

## Conclusion

In conclusion, our study contributes to the growing body of evidence on the impact of COVID-19 on the digestive system. Specifically, there are continuing risks of FGID, GERD, NAFLD and pancreatic diseases requiring long-term follow-up and further attention. Our findings highlight the need for long-term care and management of COVID-19 patients to monitor potential post-acute complications of digestive system.

## Data Availability

All data produced in the present study are available upon reasonable request to the authors

## Acknowledgement

The authors thank the UK Biobank for the access of data, and this research has been performed under approval (Application Number 83339).

## Author Contributions

YYM, LJZ, RW, and WYD contributed equally to this work. HC, WHS, CYD, and FWL are senior and corresponding authors who also contributed equally to this study. YYM, LJZ, WHS, and HC contributed to data extraction, data analyses, and manuscript drafting. YYM, LJZ, RW and WYD contributed to data interpretation and manuscript drafting. RJZ, RJ, DLL, HHW, ZWZ, QY, and JWL contributed to manuscript drafting. HC, WHS, CYD, and FWL contributed to study design, data interpretation, and final approval of the manuscript. The corresponding author attests that all listed authors meet authorship criteria and that no others meeting the criteria have been omitted.

## Abbreviations and Acronyms

(BMI): body mass index
(COPD): chronic obstructive pulmonary disease
(CI): confidence intervals
(COVID-19): coronavirus disease 2019
(DAG): directed acyclic graph
(HR): hazard ratios
(IPTW): propensity score matching
(SARS-CoV-2): severe acute respiratory syndrome coronavirus 2
(SD): standard deviation
(FGID): functional gastrointestinal disorders
(IBS): irritable bowel syndrome
(PUD): peptic ulcer disease
(IBD): inflammatory bowel disease
(CD): Crohn’s disease
(UC): ulcerative colitis
(NAFLD): non-alcohol fatty liver disease
(SMD): standardized mean difference

## Figure Legends

**Supplementary Figure 1. Directed Acyclic Graphs (DAG) for covariate selection**.

DAG showing the relationships between the study variables.

**Supplementary Figure 2. Flow chart of eligible participants selection**.

**Supplementary Figure 3. Hazard ratio of digestive outcomes in COVID-19 group and the contemporary control by severity of COVID-19**.

HR: hazard ratios; CI: confidence interval;

Outcomes were ascertained 30 days after the COVID-19-positive test until the end of follow-up. Weighted HRs after IPTW and 95% CIs are presented.

