## Supplementary materials for "Risks of digestive diseases in long COVID: Evidence from a large-scale cohort study"

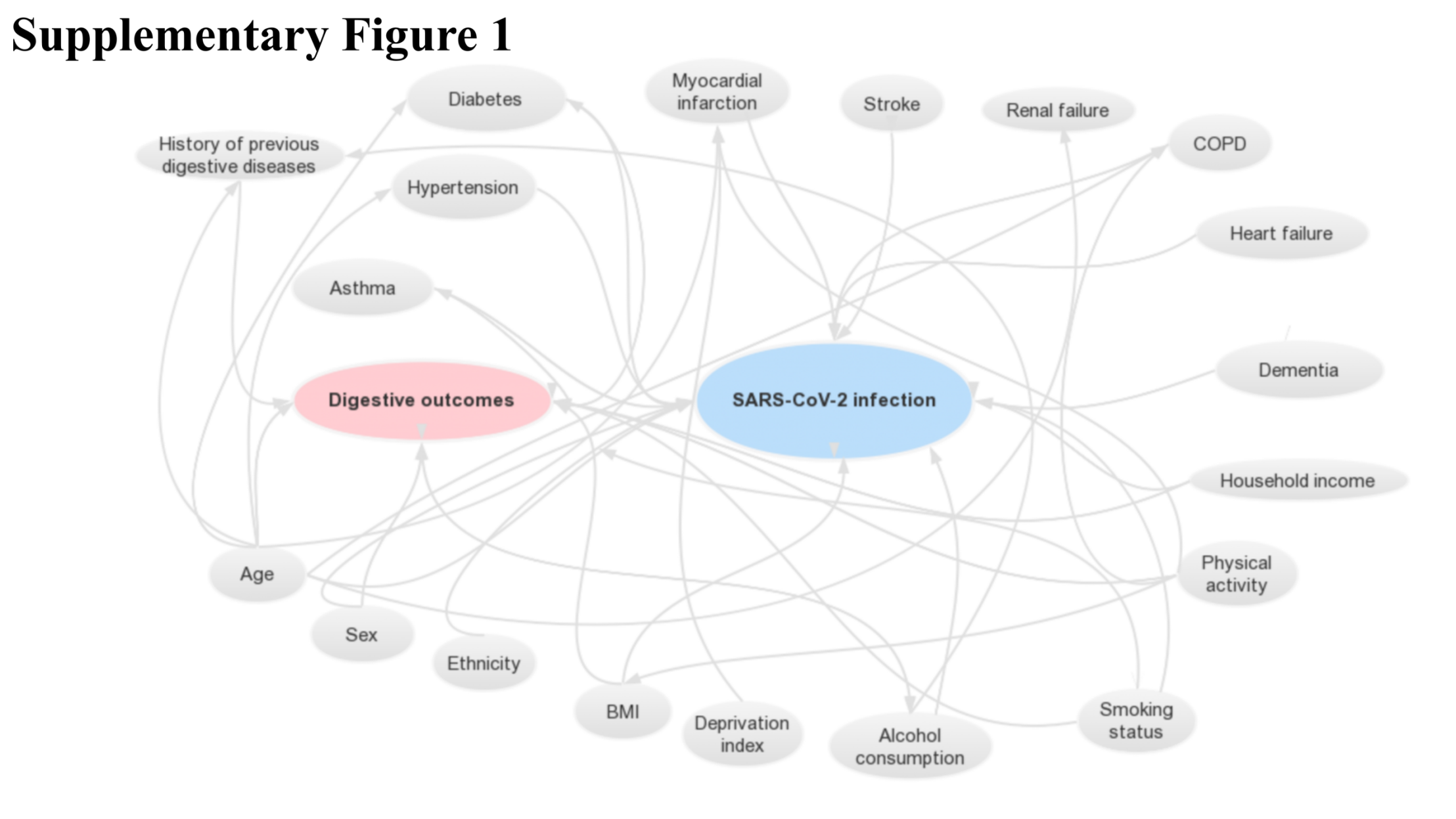

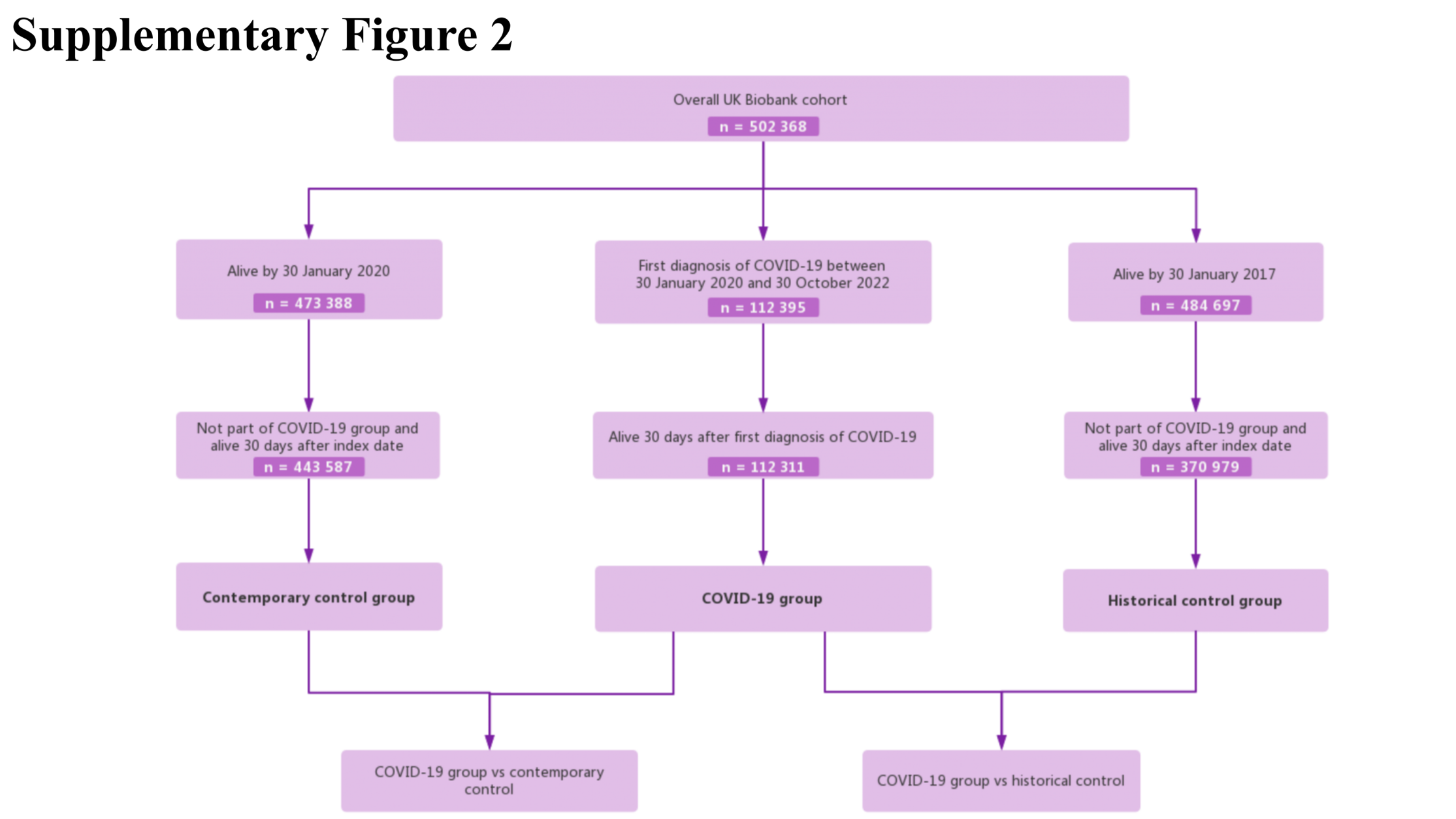

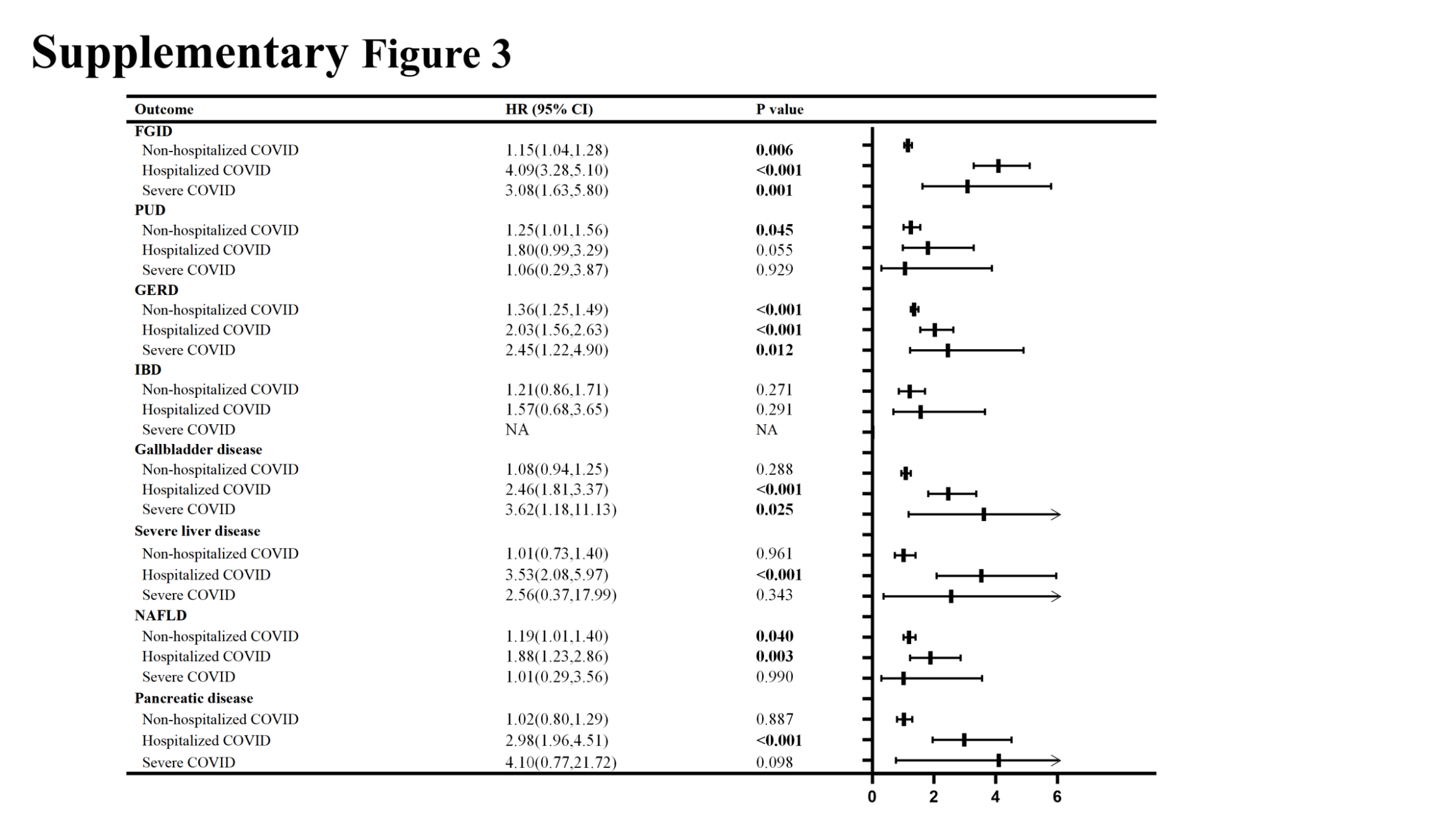
Supplementary Tables

Supplementary Table 1. Outcome ascertainment.

Supplementary Table 2. The numbers (percentages) of participants with missing covariates.

Supplementary Table 3. Baseline characteristics of COVID-19 group and contemporary controls before weighting

Supplementary Table 4. Hazard ratio of digestive outcomes in COVID-19 group and the contemporary control at different follow-up times

Supplementary Table 5. Baseline characteristics of COVID-19 group and contemporary controls by severity of COVID-19 before weighting

Supplementary Table 6. Baseline characteristics of COVID-19 group and contemporary controls by severity of COVID-19 after weighting

Supplementary Table 7. Baseline characteristics of COVID-19 group and contemporary controls in the sensitive analysis restricting to the period before vaccination was available before weighting.

Supplementary Table 8. Baseline characteristics of COVID-19 group and contemporary controls in the sensitive analysis restricting to the period before vaccination was available after weighting.

Supplementary Table 9. Hazard ratio of digestive outcomes in COVID-19 group and contemporary and historical controls in subgroups in the sensitive analysis restricting to the period before vaccination was available.

Supplementary Table 10. Hazard ratio of digestive outcomes in COVID-19 group and contemporary and historical controls in subgroups

Supplementary Table 11. Hazard ratio of digestive outcomes in COVID-19 group, the contemporary and historical control by sex

Supplementary Table 12. Baseline characteristics of COVID-19 group and historical controls before weighting

Supplementary Table 13. Baseline characteristics of COVID-19 group and historical controls after weighting

Supplementary Table 14. Baseline characteristics of COVID-19 group and historical controls by severity of COVID-19 before weighting

Supplementary Table 15. Baseline characteristics of COVID-19 group and historical controls by severity of COVID-19 after weighting

Supplementary Table 16. Hazard ratio of digestive outcomes in COVID-19 group and the historical control by severity of COVID-19.

Supplementary Table 17. Baseline characteristics of COVID-19 group and histotical controls in the sensitive analysis restricting to the period before vaccination was available before weighting.

Supplementary Table 18. Baseline characteristics of COVID-19 group and historical controls in the sensitive analysis restricting to the period before vaccination was available after weighting.

Supplementary Table 1. Outcome ascertainment.

| Outcome | ICD-10 |
| --- | --- |
| FGID |  |
| Dyspepsia | K30 |
| Irritable bowel syndrome | K58 |
| Constipation | K590 |
| PUD |  |
| Gastric ulcer | K25 |
| Duodenal ulcer | K26 |
| Other PUD | K27 |
| GERD | K21 |
| IBD |  |
| CD | K50 |
| UC | K51 |
| Gallbladder disease |  |
| Cholelithiasis | K80 |
| Cholecystitis | K81 |
| Severe liver disease |  |
| Liver failure | K704, K721, K729 |
| Hepatic sclerosis or cirrhosis | K703, K741, K742, K746 |
| Complication of liver diseases | I850, I859, K766, K767 |
| NAFLD | K758, K759, K760 |
| Pancreatic disease |  |
| Acute pancreatitis | K85 |
| Chronic pancreatitis | K861 |
| Pancreatic cyst | K862, K863 |
| Other diseases of pancreatic | K868, K869 |

Supplementary Table 2. The numbers (percentages) of participants with missing covariates

| Covariates | N | % |
| --- | --- | --- |
| Ethnicity | 2 777 | 0.55% |
| Household income | 225 | 0.04% |
| Deprivation index | 624 | 0.12% |
| BMI | 3 105 | 0.62% |
| Alcohol consumption | 1 501 | 0.30% |
| Smoking status | 2 949 | 0.59% |
| Physical activity | 100 103 | 19.93% |

BMI: body mass index; MET: metabolic equivalent of task; COPD: chronic obstructive pulmonary disease; SD: standard deviation

Supplementary Table 3. Baseline characteristics of COVID-19 group and contemporary controls before weighting

| Characteristics | COVID-19 group  (n = 112 311) | Contemporary controls  (n = 359 671) | SMD |
| --- | --- | --- | --- |
| Age, mean(SD), years | 54.43(8.19) | 56.79(7.97) | 0.293 |
| Sex, female, n(%) | 61 495(54.8) | 199 368(55.4) | 0.014 |
| Ethnicity, White, n(%) | 106 315(94.7) | 339 676(94.4) | 0.010 |
| Household income |  |  | 0.189 |
| <18 000, n(%) | 19 465(17.3) | 82 983(23.1) |  |
| 18 000-30 999, n(%) | 26 102(23.2) | 94 121(26.2) |  |
| 31 000-51 999, n(%) | 31 853(28.4) | 92 659(25.8) |  |
| 52 000-100 000, n(%) | 27 297(24.3) | 70 470(19.6) |  |
| >100 000, n(%) | 7 594(6.8) | 19 438(5.4) |  |
| Deprivation index, mean(SD) | -1.38(3.00) | -1.32(3.09) | 0.023 |
| BMI, mean(SD), kg/m^2^ | 27.47(4.82) | 27.36(4.74) | 0.025 |
| Alcohol consumption |  |  | 0.099 |
| Daily or almost daily, n(%) | 21 925(19.5) | 73 209(20.4) |  |
| Three or four times a week, n(%) | 28 027(25.0) | 81 886(22.8) |  |
| Once or twice a week, n(%) | 30 734(27.4) | 91 957(25.6) |  |
| One to three times a month, n(%) | 12 790(11.4) | 40 314(11.2) |  |
| Special occasions only or never, n(%) | 11 298(10.1) | 42 710(11.9) |  |
| Never, n(%) | 7 537(6.7) | 29 595(8.2) |  |
| Smoking status |  |  | 0.053 |
| Never smoker, n(%) | 63 505(56.5) | 199 721(55.5) |  |
| Previous smoker, n(%) | 38 875(34.6) | 122 502(34.1) |  |
| Current smoker, n(%) | 9 931(8.8) | 37 448(10.4) |  |
| Physical activity, mean(SD), MET minutes/week | 2536.46(2603.69) | 2686.42(2739.02) | 0.056 |
| Comorbidities |  |  |  |
| Hypertension, n(%) | 38 023(33.9) | 132 105(36.7) | 0.060 |
| Diabetes, n(%) | 8 384(7.5) | 27 531(7.7) | 0.007 |
| Renal failure, n(%) | 4 646(4.1) | 15 075(4.2) | 0.003 |
| Myocardial infarction, n(%) | 4 557(4.1) | 15 720(4.4) | 0.016 |
| Stroke, n(%) | 2 706(2.4) | 9 577(2.7) | 0.016 |
| COPD, n(%) | 1 7231(15.3) | 48 079(13.4) | 0.056 |
| Asthma, n(%) | 4 377(3.9) | 14 699(4.1) | 0.010 |
| Heart failure, n(%) | 2 356(2.1) | 7 888(2.2) | 0.007 |
| Dementia, n(%) | 1 060(0.9) | 2 322(0.6) | 0.034 |
| History of previous digestive diseases, n(%) | 37 375(33.3) | 116 937(32.5) | 0.016 |

SMD: standard mean difference; BMI: body mass index; MET: metabolic equivalent of task; COPD: chronic obstructive pulmonary disease; SD: standard deviation

Supplementary Table 4. Hazard ratio of digestive outcomes in COVID-19 group and the contemporary control at different follow-up times

| Outcome | 0 to 6-month follow-up | | 6 to 12-month follow-up | | More than 12-month follow up | |
| --- | --- | --- | --- | --- | --- | --- |
|  | HR (95% CI) | P value | HR (95% CI) | P value | HR (95% CI) | P value |
| FGID | 1.68(1.50,1.88) | **<0.001** | 1.18(0.98,1.42) | 0.078 | 1.40(1.13,1.73) | **0.002** |
| PUD | 1.34(1.02,1.76) | **0.036** | 1.05(0.70,1.57) | 0.823 | 1.55(0.96,2.50) | 0.071 |
| GERD | 1.55(1.39,1.73) | **<0.001** | 1.24(1.05,1.46) | **0.010** | 1.65(1.36,1.99) | **<0.001** |
| IBD | 1.46(0.99,2.14) | 0.055 | 1.42(0.74,2.72) | 0.291 | 1.33(0.56,3.19) | 0.518 |
| Gallbladder disease | 1.29(1.09,1.54) | **0.003** | 1.26(0.98,1.62) | 0.071 | 1.27(0.92,1.75) | 0.151 |
| Severe liver disease | 1.33(0.92,1.93) | 0.132 | 1.57(0.95,2.61) | 0.080 | 1.70(0.95,3.05) | 0.073 |
| NAFLD | 1.53(1.26,1.85) | **<0.001** | 0.96(0.70,1.32) | 0.806 | 1.54(1.11,2.12) | **0.009** |
| Pancreatic disease | 1.58(1.22,2.05) | **0.001** | 0.96(0.61,1.49) | 0.850 | 1.73(1.13,2.65) | **0.011** |

HR: hazard ratio; CI: confidence interval;

Weighted HRs after IPTW and 95% CIs are presented.

Supplementary Table 5. Baseline characteristics of COVID-19, contemporary controls by severity of COVID-19 before weighting

| Characteristics | Non-hospitalized COVID  (n= 104 201) | Hospitalized COVID  (n= 7 523) | Severe COVID  (n= 588) | Contemporary controls  (n = 359 671) | SMD | | |
| --- | --- | --- | --- | --- | --- | --- | --- |
|  |  |  |  |  | Non-hospitalized COVID and contemporary controls | Hospitalized COVID and contemporary controls | Severe COVID and contemporary controls |
| Age, mean(SD), years | 54.02(8.09) | 59.84(7.55) | 57.78(7.99) | 56.79(7.97) | 0.346 | 0.393 | 0.124 |
| Sex, female, n(%) | 57 959(55.6) | 3 324(44.2) | 212(36.1) | 199 368(55.4) | 0.004 | 0.226 | 0.397 |
| Ethnicity, White, n(%) | 98 873(94.9) | 6 951(92.4) | 492(83.7) | 339 676(94.4) | 0.020 | 0.083 | 0.350 |
| Household income |  |  |  |  | 0.232 | 0.335 | 0.391 |
| <18 000, n(%) | 16 497(15.8) | 2 741(36.4) | 227(38.6) | 82 983(23.1) |  |  |  |
| 18 000-30 999, n(%) | 23 945(23.0) | 2 003(26.6) | 154(26.2) | 94 121(26.2) |  |  |  |
| 31 000-51 999, n(%) | 30 165(28.9) | 1 567(20.8) | 122(20.7) | 92 659(25.8) |  |  |  |
| 52 000-100 000, n(%) | 26 266(25.2) | 961(12.8) | 70(11.9) | 70 470(19.6) |  |  |  |
| >100 000, n(%) | 7 328(7.0) | 251(3.3) | 15(2.6) | 19 438(5.4) |  |  |  |
| Deprivation index, mean(SD) | -1.46(2.95) | -0.43(3.40) | 0.16(3.54) | -1.32(3.09) | 0.049 | 0.273 | 0.443 |
| BMI, mean(SD), kg/m^2^ | 27.33(4.72) | 29.23(5.63) | 30.61(5.78) | 27.36(4.74) | 0.006 | 0.360 | 0.615 |
| Alcohol consumption |  |  |  |  | 0.124 | 0.209 | 0.316 |
| Daily or almost daily, n(%) | 20 416(19.6) | 1 414(18.8) | 96(16.3) | 73 209(20.4) |  |  |  |
| Three or four times a week, n(%) | 26 582(25.5) | 1353(18.0) | 92(15.6) | 81 886(22.8) |  |  |  |
| Once or twice a week, n(%) | 28 762(27.6) | 1 825(24.3) | 147(25.0) | 91 957(25.6) |  |  |  |
| One to three times a month, n(%) | 11 927(11.4) | 799(10.6) | 64(10.9) | 40 314(11.2) |  |  |  |
| Special occasions only or never, n(%) | 10 034(9.6) | 1 168(15.5) | 96(16.3) | 42 710(11.9) |  |  |  |
| Never, n(%) | 6 480(6.2) | 964(12.8) | 93(15.8) | 29 595(8.2) |  |  |  |
| Smoking status |  |  |  |  | 0.070 | 0.228 | 0.263 |
| Never smoker, n(%) | 59 921(57.5) | 3 335(44.3) | 250(42.5) | 199 721(55.5) |  |  |  |
| Previous smoker, n(%) | 35 500(34.1) | 3 115(41.4) | 260(44.2) | 122 502(34.1) |  |  |  |
| Current smoker, n(%) | 8 780(8.4) | 1 073(14.3) | 78(13.3) | 37 448(10.4) |  |  |  |
| Physical activity, mean(SD), MET minutes/week | 2529.22(2585.74) | 2623.26(2813.78) | 2707.49(2935.05) | 2686.42(2739.02) | 0.059 | 0.023 | 0.007 |
| Comorbidities |  |  |  |  |  |  |  |
| Hypertension, n(%) | 33 161(31.8) | 4 523(60.1) | 339(57.7) | 132 105(36.7) | 0.103 | 0.481 | 0.429 |
| Diabetes, n(%) | 6 579(6.3) | 1 678(22.3) | 127(21.6) | 27 531(7.7) | 0.053 | 0.419 | 0.403 |
| Renal failure, n(%) | 3 515(3.4) | 1 050(14.0) | 81(13.8) | 15 075(4.2) | 0.043 | 0.345 | 0.34 |
| Myocardial infarction, n(%) | 3 599(3.5) | 890(11.8) | 68(11.6) | 15 720(4.4) | 0.047 | 0.276 | 0.268 |
| Stroke, n(%) | 2 056(2.0) | 623(8.3) | 27(4.6) | 9 577(2.7) | 0.046 | 0.249 | 0.103 |
| COPD, n(%) | 3 174(3.0) | 1 123(14.9) | 80(13.6) | 48 079(13.4) | 0.056 | 0.376 | 0.340 |
| Asthma, n(%) | 15 561(14.9) | 1 534(20.4) | 136(23.1) | 14 699(4.1) | 0.045 | 0.188 | 0.255 |
| Heart failure, n(%) | 1 607(1.5) | 714(9.5) | 35(6.0) | 7 888(2.2) | 0.048 | 0.315 | 0.191 |
| Dementia, n(%) | 732(0.7) | 321(4.3) | 7(1.2) | 2 322(0.6) | 0.007 | 0.236 | 0.057 |
| History of previous digestive diseases, n(%) | 33 024(31.7) | 4 085(54.3) | 266(45.2) | 116 937(32.5) | 0.018 | 0.451 | 0.263 |

SMD: standard mean difference; BMI: body mass index; MET: metabolic equivalent of task; COPD: chronic obstructive pulmonary disease; SD: standard deviation

Supplementary Table 6. Baseline characteristics of COVID-19, contemporary controls by severity of COVID-19 after weighting

| Characteristics | Non-hospitalized COVID  (n= 104 201) | Hospitalized COVID  (n= 7 523) | Severe COVID  (n= 588) | Contemporary controls  (n = 359 671) | SMD | | |
| --- | --- | --- | --- | --- | --- | --- | --- |
|  |  |  |  |  | Non-hospitalized COVID and contemporary controls | Hospitalized COVID and contemporary controls | Severe COVID and contemporary controls |
| Age, mean(SD), years | 56.1(8.0) | 56.8(8.3) | 57.4(7.7) | 56.79(7.97) | 0.003 | 0.011 | 0.078 |
| Sex, female, n(%) | 57 415(55.1) | 4 093(54.4) | 297(50.5) | 199 368(55.4) | 0.007 | 0.016 | 0.098 |
| Ethnicity, White, n(%) | 98 574(94.6) | 7 072(94) | 549(93.4) | 339 676(94.4) | 0.002 | 0.015 | 0.042 |
| Household income |  |  |  |  | 0.005 | 0.033 | 0.079 |
| <18 000, n(%) | 22 195(21.3) | 1 813(24.1) | 148(25.2) | 82 983(23.1) |  |  |  |
| 18 000-30 999, n(%) | 26 467(25.4) | 1 933(25.7) | 163(27.8) | 94 121(26.2) |  |  |  |
| 31 000-51 999, n(%) | 27 613(26.5) | 1 994(26.5) | 143(24.4) | 92 659(25.8) |  |  |  |
| 52 000-100 000, n(%) | 21 882(21) | 1 407(18.7) | 102(17.3) | 70 470(19.6) |  |  |  |
| >100 000, n(%) | 6 044(5.8) | 384(5.1) | 31(5.3) | 19 438(5.4) |  |  |  |
| Deprivation index, mean(SD) | -1.4(3.0) | -1.3(3.1) | -1.1(3.2) | -1.32(3.09) | 0.006 | 0.014 | 0.059 |
| BMI, mean(SD), kg/m^2^ | 27.3(4.7) | 27.6(5.0) | 28.3(4.7) | 27.36(4.74) | 0.004 | 0.046 | 0.193 |
| Alcohol consumption |  |  |  |  | 0.007 | 0.044 | 0.148 |
| Daily or almost daily, n(%) | 21 257(20.4) | 1 535(20.4) | 107(18.2) | 73 209(20.4) |  |  |  |
| Three or four times a week, n(%) | 24 487(23.5) | 1 625(21.6) | 115(19.6) | 81 886(22.8) |  |  |  |
| Once or twice a week, n(%) | 27 092(26.0) | 1 866(24.8) | 152(25.9) | 91 957(25.6) |  |  |  |
| One to three times a month, n(%) | 11 671(11.2) | 895(11.9) | 78(13.3) | 40 314(11.2) |  |  |  |
| Special occasions only or never, n(%) | 11 671(11.2) | 963(12.8) | 66(11.3) | 42 710(11.9) |  |  |  |
| Never, n(%) | 8 128(7.8) | 639(8.5) | 68(11.6) | 29 595(8.2) |  |  |  |
| Smoking status |  |  |  |  | 0.005 | 0.023 | 0.076 |
| Never smoker, n(%) | 58 353(56) | 4 093(54.4) | 329(55.9) | 199 721(55.5) |  |  |  |
| Previous smoker, n(%) | 35 533(34.1) | 2 603(34.6) | 186(31.6) | 122 502(34.1) |  |  |  |
| Current smoker, n(%) | 10 212(9.8) | 835(11.1) | 74(12.5) | 37 448(10.4) |  |  |  |
| Physical activity, mean(SD), MET minutes/week | 2644.4(2696.7) | 2649.8(2706.1) | 2688.7(2790.6) | 2686.42(2739.02) | 0.002 | 0.013 | 0.001 |
| Comorbidities |  |  |  |  |  |  |  |
| Hypertension, n(%) | 37 096(35.6) | 2 836(37.7) | 253(43.1) | 132 105(36.7) | 0.001 | 0.010 | 0.130 |
| Diabetes, n(%) | 7 607(7.3) | 639(8.5) | 62(10.5) | 27 531(7.7) | <0.001 | 0.018 | 0.097 |
| Renal failure, n(%) | 4 168(4) | 361(4.8) | 31(5.3) | 15 075(4.2) | <0.001 | 0.019 | 0.053 |
| Myocardial infarction, n(%) | 4 376(4.2) | 354(4.7) | 35(5.9) | 15 720(4.4) | <0.001 | 0.009 | 0.068 |
| Stroke, n(%) | 2 605(2.5) | 233(3.1) | 19(3.2) | 9 577(2.7) | 0.002 | 0.016 | 0.032 |
| COPD, n(%) | 3 960(3.8) | 617(8.2) | 55(9.3) | 48 079(13.4) | 0.007 | 0.165 | 0.209 |
| Asthma, n(%) | 14 276(13.7) | 1 038(13.8) | 96(16.4) | 14 699(4.1) | 0.001 | 0.010 | 0.085 |
| Heart failure, n(%) | 1 980(1.9) | 323(4.3) | 22(3.7) | 7 888(2.2) | 0.011 | 0.112 | 0.088 |
| Dementia, n(%) | 729(0.7) | 75(1.0) | 2(0.4) | 2 322(0.6) | 0.004 | 0.029 | 0.038 |
| History of previous digestive diseases, n(%) | 33 657(32.3) | 2 580(34.3) | 223(38) | 116 937(32.5) | <0.001 | 0.028 | 0.115 |

SMD: standard mean difference; BMI: body mass index; MET: metabolic equivalent of task; COPD: chronic obstructive pulmonary disease; SD: standard deviation

Supplementary Table 7. Baseline characteristics of COVID-19 group and contemporary controls in the sensitive analysis restricting to the period before vaccination was available before weighting.

| Characteristics | COVID-19 group  (n = 8 431) | Contemporary controls  (n = 359 671) | SMD |
| --- | --- | --- | --- |
| Age, mean(SD), years | 54.39(8.82) | 56.79(7.97) | 0.286 |
| Sex, female, n(%) | 4 280(50.8) | 199 368(55.4) | 0.094 |
| Ethnicity, White, n(%) | 7 771(92.2) | 339 676(94.4) | 0.091 |
| Household income |  |  | 0.073 |
| <18 000, n(%) | 2 147(25.5) | 82 983(23.1) |  |
| 18 000-30 999, n(%) | 2 119(25.1) | 94 121(26.2) |  |
| 31 000-51 999, n(%) | 2 236(26.5) | 92 659(25.8) |  |
| 52 000-100 000, n(%) | 1 546(18.3) | 70 470(19.6) |  |
| >100 000, n(%) | 383(4.5) | 19 438(5.4) |  |
| Deprivation index, mean(SD) | -0.73(3.27) | -1.32(3.09) | 0.184 |
| BMI, mean(SD), kg/m^2^ | 28.43(5.07) | 27.36(4.74) | 0.218 |
| Alcohol consumption |  |  | 0.114 |
| Daily or almost daily, n(%) | 1 408(16.7) | 73 209(20.4) |  |
| Three or four times a week, n(%) | 1 843(21.9) | 81 886(22.8) |  |
| Once or twice a week, n(%) | 2 394(28.4) | 91 957(25.6) |  |
| One to three times a month, n(%) | 965(11.4) | 40 314(11.2) |  |
| Special occasions only or never, n(%) | 1 004(11.9) | 42 710(11.9) |  |
| Never, n(%) | 817(9.7) | 29 595(8.2) |  |
| Smoking status |  |  | 0.086 |
| Never smoker, n(%) | 4 322(51.3) | 199 721(55.5) |  |
| Previous smoker, n(%) | 3 115(36.9) | 122 502(34.1) |  |
| Current smoker, n(%) | 994(11.8) | 37 448(10.4) |  |
| Physical activity, mean(SD), MET minutes/week | 2734.76(2828.44) | 2686.42(2739.02) | 0.017 |
| Comorbidities |  |  |  |
| Hypertension, n(%) | 3 411(40.5) | 132 105(36.7) | 0.077 |
| Diabetes, n(%) | 1 005(11.9) | 27 531(7.7) | 0.144 |
| Renal failure, n(%) | 573(6.8) | 15 075(4.2) | 0.115 |
| Myocardial infarction, n(%) | 547(6.5) | 15 720(4.4) | 0.094 |
| Stroke, n(%) | 398(4.7) | 9 577(2.7) | 0.109 |
| COPD, n(%) | 616(7.3) | 14 699(4.1) | 0.139 |
| Asthma, n(%) | 1 382(16.4) | 48 079(13.4) | 0.085 |
| Heart failure, n(%) | 385(4.6) | 7 888(2.2) | 0.132 |
| Dementia, n(%) | 345(4.1) | 2 322(0.6) | 0.228 |
| History of previous digestive diseases, n(%) | 3 286(39.0) | 116 937(32.5) | 0.135 |

SMD: standard mean difference; BMI: body mass index; MET: metabolic equivalent of task; COPD: chronic obstructive pulmonary disease; SD: standard deviation

Supplementary Table 8. Baseline characteristics of COVID-19 group and contemporary controls in the sensitive analysis restricting to the period before vaccination was available after weighting.

| Characteristics | COVID-19 group  (n = 8 431) | Contemporary controls  (n = 359 671) | SMD |
| --- | --- | --- | --- |
| Age, mean(SD), years | 56.7(8.5) | 56.7(8.0) | 0.001 |
| Sex, female, n(%) | 4 519(53.6) | 198 898(55.3) | 0.034 |
| Ethnicity, White, n(%) | 7 934(94.1) | 339 529(94.4) | 0.010 |
| Household income |  |  | 0.030 |
| <18 000, n(%) | 2 049(24.3) | 83 084(23.1) |  |
| 18 000-30 999, n(%) | 2 167(25.7) | 93 874(26.1) |  |
| 31 000-51 999, n(%) | 2 150(25.5) | 92 795(25.8) |  |
| 52 000-100 000, n(%) | 1 602(19) | 70 496(19.6) |  |
| >100 000, n(%) | 464(5.5) | 19 422(5.4) |  |
| Deprivation index, mean(SD) | -1.2(3.1) | -1.3(3.1) | 0.034 |
| BMI, mean(SD), kg/m^2^ | 27.6(4.6) | 27.4(4.8) | 0.042 |
| Alcohol consumption |  |  | 0.022 |
| Daily or almost daily, n(%) | 1 728(20.5) | 73 013(20.3) |  |
| Three or four times a week, n(%) | 1 889(22.4) | 81 645(22.7) |  |
| Once or twice a week, n(%) | 2 125(25.2) | 92 076(25.6) |  |
| One to three times a month, n(%) | 936(11.1) | 40 283(11.2) |  |
| Special occasions only or never, n(%) | 1 012(12) | 42 801(11.9) |  |
| Never, n(%) | 742(8.8) | 29 853(8.3) |  |
| Smoking status |  |  | 0.040 |
| Never smoker, n(%) | 4 553(54) | 199 258(55.4) |  |
| Previous smoker, n(%) | 2 900(34.4) | 122 648(34.1) |  |
| Current smoker, n(%) | 978(11.6) | 37 406(10.4) |  |
| Physical activity, mean(SD), MET minutes/week | 2687.5 (2742.4) | 2676.1 (2715.0) | 0.004 |
| Comorbidities |  |  |  |
| Hypertension, n(%) | 3 238(38.4) | 132 359(36.8) | 0.032 |
| Diabetes, n(%) | 700(8.3) | 28 054(7.8) | 0.019 |
| Renal failure, n(%) | 363(4.3) | 15 466(4.3) | 0.003 |
| Myocardial infarction, n(%) | 379(4.5) | 15 826(4.4) | 0.005 |
| Stroke, n(%) | 261(3.1) | 9 711(2.7) | 0.026 |
| COPD, n(%) | 582(6.9) | 14 747(4.1) | **0.121** |
| Asthma, n(%) | 1 155(13.7) | 48 196(13.4) | 0.009 |
| Heart failure, n(%) | 304(3.6) | 7913(2.2) | 0.083 |
| Dementia, n(%) | 67(0.8) | 2518(0.7) | 0.012 |
| History of previous digestive diseases, n(%) | 3 263(38.7) | 117 253(32.6) | 0.128 |

SMD: standard mean difference; BMI: body mass index; MET: metabolic equivalent of task; COPD: chronic obstructive pulmonary disease; SD: standard deviation

Supplementary Table 9. Hazard ratio of digestive outcomes in COVID-19 group and contemporary and historical controls in subgroups in the sensitive analysis restricting to the period before vaccination was available.

| Outcome | COVID-19 vs Contemporary control | | COVID-19 vs Historical control | |
| --- | --- | --- | --- | --- |
|  | HR (95% CI) | P value | HR (95% CI) | P value |
| FGID | 2.12(1.67,2.69) | **<0.001** | 1.63(1.36,1.95) | **<0.001** |
| PUD | 1.16(0.61,2.22) | 0.654 | 0.88(0.56,1.41) | 0.604 |
| GERD | 1.49(1.14,1.93) | **0.003** | 1.42(1.19,1.68) | **<0.001** |
| IBD | 2.21(1.13,4.35) | 0.021 | 1.39(0.76,2.55) | 0.283 |
| Severe liver disease | 1.64(1.15,2.34) | **0.007** | 1.42(1.09,1.85) | **0.010** |
| NAFLD | 1.33(0.84,2.12) | 0.222 | 2.10(1.56,2.84) | **<0.001** |
| Gallbladder disease | 2.08(1.18,3.67) | **0.012** | 2.24(1.48,3.39) | **<0.001** |
| Pancreatic disease | 2.12(1.67,2.69) | **<0.001** | 1.63(1.36,1.95) | **<0.001** |

HR: hazard ratio; CI: confidence interval;

Outcomes were ascertained 30 days after the COVID-19-positive test until the end of follow-up. Weighted HRs after IPTW and 95% CIs are presented.

Supplement Table 10. Hazard ratio of digestive outcomes compared with contemporary and historical controls in subgroups

| **Outcome** | **Subgroup** | **COVID-19 vs Contemporary control** | | | **COVID-19 vs Historical control** | | |
| --- | --- | --- | --- | --- | --- | --- | --- |
|  |  | **HR** | **95% CI** | **Pi** | **HR** | **95% CI** | **Pi** |
|  | **Age** |  |  |  |  |  |  |
| **FGID** | Age<60 | 1.44 | 1.27,1.65 | 0.392 | 1.31 | 1.16,1.48 | <0.001 |
|  | Age>=60 | 1.56 | 1.38,1.75 |  | 2.01 | 1.79,2.26 |  |
| **PUD** | Age<60 | 1.27 | 0.96,1.69 | 0.891 | 1.02 | 0.78,1.33 | 0.933 |
|  | Age>=60 | 1.30 | 0.97,1.74 |  | 1.00 | 0.76,1.30 |  |
| **GERD** | Age<60 | 1.39 | 1.25,1.55 | 0.170 | 1.37 | 1.24,1.51 | 0.838 |
|  | Age>=60 | 1.57 | 1.38,1.78 |  | 1.44 | 1.28,1.62 |  |
| **IBD** | Age<60 | 1.04 | 0.69,1.57 | 0.046 | 0.95 | 0.65,1.39 | 0.127 |
|  | Age>=60 | 1.95 | 1.24,3.09 |  | 1.68 | 1.09,2.57 |  |
| **Severe liver disease** | Age<60 | 1.31 | 0.91,1.88 | 0.381 | 1.11 | 0.95,1.30 | 0.162 |
|  | Age>=60 | 1.66 | 1.13,2.44 |  | 1.44 | 1.19,1.74 |  |
| **NAFLD** | Age<60 | 1.21 | 1.00,1.46 | 0.113 | 1.85 | 1.31,2.61 | 0.568 |
|  | Age>=60 | 1.53 | 1.22,1.92 |  | 2.46 | 1.72,3.51 |  |
| **Gallbladder disease** | Age<60 | 1.27 | 1.07,1.50 | 0.688 | 2.57 | 2.10,3.14 | 0.062 |
|  | Age>=60 | 1.34 | 1.10,1.62 |  | 3.41 | 2.66,4.36 |  |
| **Pancreatic disease** | Age<60 | 1.53 | 1.15,2.03 | 0.632 | 1.83 | 1.40,2.41 | 0.896 |
|  | Age>=60 | 1.41 | 1.06,1.86 |  | 2.52 | 1.85,3.44 |  |
|  | **Ethnicity** |  |  |  |  |  |  |
| **FGID** | White | 1.47 | 1.34,1.61 | 0.641 | 1.64 | 1.50,1.79 | 0.720 |
|  | Other | 1.63 | 1.12,2.37 |  | 1.45 | 1.06,2.00 |  |
| **PUD** | White | 1.26 | 1.02,1.55 | 0.662 | 0.98 | 0.80,1.19 | 0.879 |
|  | Other | 1.42 | 0.66,3.05 |  | 0.98 | 0.49,1.95 |  |
| **GERD** | White | 1.48 | 1.36,1.61 | 0.225 | 1.39 | 1.28,1.50 | 0.416 |
|  | Other | 1.18 | 0.85,1.65 |  | 1.25 | 0.92,1.71 |  |
| **IBD** | White | 1.45 | 1.06,1.99 | 0.255 | 1.31 | 0.98,1.77 | 0.093 |
|  | Other | 0.56 | 0.11,2.74 |  | 0.43 | 0.10,1.84 |  |
| **Severe liver disease** | White | 1.53 | 1.17,2.00 | 0.237 | 1.22 | 1.07,1.39 | 0.238 |
|  | Other | 0.66 | 0.15,2.92 |  | 1.91 | 1.15,3.17 |  |
| **NAFLD** | White | 1.31 | 1.12,1.53 | 0.510 | 2.21 | 1.71,2.84 | 0.364 |
|  | Other | 1.56 | 0.92,2.63 |  | 0.91 | 0.22,3.82 |  |
| **Gallbladder disease** | White | 1.27 | 1.11,1.45 | 0.304 | 2.82 | 2.40,3.33 | 0.420 |
|  | Other | 1.73 | 0.99,3.00 |  | 3.49 | 2.05,5.94 |  |
| **Pancreatic disease** | White | 1.41 | 1.15,1.74 | 0.608 | 2.11 | 1.69,2.62 | 0.771 |
|  | Other | 1.77 | 0.77,4.09 |  | 2.68 | 1.18,6.07 |  |
|  | **Smoke status** |  |  |  |  |  |  |
| **FGID** | Never smoke | 1.38 | 1.21,1.57 |  | 1.51 | 1.33,1.71 |  |
|  | Previous smoke | 1.52 | 1.33,1.75 | 0.298 | 1.75 | 1.53,2.00 | 0.334 |
|  | Current smoke | 1.68 | 1.31,2.17 | 0.171 | 1.65 | 1.31,2.08 | 0.237 |
| **PUD** | Never smoke | 1.22 | 0.90,1.65 |  | 1.00 | 0.75,1.33 |  |
|  | Previous smoke | 1.41 | 1.03,1.93 | 0.506 | 1.06 | 0.79,1.41 | 0.573 |
|  | Current smoke | 1.04 | 0.56,1.93 | 0.627 | 0.73 | 0.43,1.26 | 0.389 |
| **GERD** | Never smoke | 1.41 | 1.26,1.58 |  | 1.37 | 1.23,1.52 |  |
|  | Previous smoke | 1.45 | 1.27,1.66 | 0.754 | 1.32 | 1.16,1.51 | 0.503 |
|  | Current smoke | 1.68 | 1.32,2.13 | 0.210 | 1.57 | 1.26,1.96 | 0.137 |
| **IBD** | Never smoke | 0.94 | 0.55,1.60 |  | 0.77 | 0.46,1.27 |  |
|  | Previous smoke | 1.76 | 1.13,2.76 | 0.076 | 1.58 | 1.05,2.39 | 0.038 |
|  | Current smoke | 1.64 | 0.78,3.46 | 0.226 | 1.84 | 0.92,3.64 | 0.053 |
| **Severe liver disease** | Never smoke | 1.31 | 0.86,2.01 |  | 1.30 | 1.09,1.55 |  |
|  | Previous smoke | 1.76 | 1.19,2.60 | 0.322 | 1.21 | 0.99,1.48 | 0.731 |
|  | Current smoke | 1.04 | 0.53,2.07 | 0.593 | 1.11 | 0.76,1.62 | 0.933 |
| **NAFLD** | Never smoke | 1.41 | 1.14,1.75 |  | 2.13 | 1.42,3.20 |  |
|  | Previous smoke | 1.24 | 0.98,1.57 | 0.466 | 2.30 | 1.60,3.29 | 0.321 |
|  | Current smoke | 1.34 | 0.90,2.00 | 0.769 | 1.50 | 0.79,2.84 | 0.352 |
| **Gallbladder disease** | Never smoke | 1.34 | 1.12,1.61 |  | 3.33 | 2.61,4.26 |  |
|  | Previous smoke | 1.23 | 1.00,1.52 | 0.575 | 2.79 | 2.18,3.58 | 0.310 |
|  | Current smoke | 1.20 | 0.82,1.75 | 0.592 | 2.12 | 1.49,3.01 | 0.512 |
| **Pancreatic disease** | Never smoke | 1.39 | 1.04,1.86 |  | 1.87 | 1.41,2.50 |  |
|  | Previous smoke | 1.53 | 1.11,2.11 | 0.695 | 2.62 | 1.80,3.80 | 0.665 |
|  | Current smoke | 1.24 | 0.73,2.11 | 0.686 | 1.84 | 1.09,3.09 | 0.803 |
|  | **Alcohol consumption** |  |  |  |  |  |  |
| **FGID** | Daily or almost daily | 1.37 | 1.12,1.67 |  | 1.88 | 1.54,2.28 |  |
|  | Three or four times a week | 1.44 | 1.19,1.74 | 0.682 | 1.48 | 1.24,1.77 | 0.979 |
|  | Once or twice a week | 1.47 | 1.23,1.76 | 0.552 | 1.51 | 1.28,1.79 | 0.759 |
|  | One to three times a month | 1.21 | 0.91,1.59 | 0.498 | 1.47 | 1.12,1.91 | 0.365 |
|  | Special occasions only | 1.52 | 1.18,1.95 | 0.486 | 1.59 | 1.25,2.02 | 0.945 |
|  | Never | 1.95 | 1.50,2.54 | 0.033 | 1.82 | 1.43,2.31 | 0.110 |
| **PUD** | Daily or almost daily | 1.28 | 0.82,1.98 |  | 0.99 | 0.68,1.44 |  |
|  | Three or four times a week | 1.27 | 0.82,1.96 | 0.916 | 1.04 | 0.69,1.58 | 0.932 |
|  | Once or twice a week | 1.12 | 0.73,1.72 | 0.596 | 0.91 | 0.60,1.37 | 0.477 |
|  | One to three times a month | 0.97 | 0.52,1.83 | 0.443 | 0.86 | 0.47,1.59 | 0.441 |
|  | Special occasions only | 0.90 | 0.48,1.67 | 0.312 | 0.62 | 0.35,1.13 | 0.175 |
|  | Never | 2.56 | 1.44,4.55 | 0.070 | 1.48 | 0.91,2.41 | 0.295 |
| **GERD** | Daily or almost daily | 1.34 | 1.09,1.63 |  | 1.25 | 1.03,1.52 |  |
|  | Three or four times a week | 1.40 | 1.18,1.67 | 0.692 | 1.44 | 1.22,1.70 | 0.160 |
|  | Once or twice a week | 1.53 | 1.31,1.79 | 0.273 | 1.43 | 1.24,1.65 | 0.119 |
|  | One to three times a month | 1.21 | 0.94,1.56 | 0.582 | 1.11 | 0.87,1.40 | 0.723 |
|  | Special occasions only | 1.44 | 1.16,1.80 | 0.589 | 1.45 | 1.18,1.80 | 0.211 |
|  | Never | 1.98 | 1.51,2.58 | 0.022 | 1.47 | 1.16,1.86 | 0.233 |
| **IBD** | Daily or almost daily | 1.35 | 0.64,2.86 |  | 1.09 | 0.54,2.21 |  |
|  | Three or four times a week | 1.29 | 0.62,2.70 | 0.957 | 1.00 | 0.51,1.97 | 0.955 |
|  | Once or twice a week | 1.66 | 0.93,2.97 | 0.655 | 1.59 | 0.94,2.70 | 0.377 |
|  | One to three times a month | 1.35 | 0.61,2.99 | 0.992 | 1.36 | 0.70,2.66 | 0.772 |
|  | Special occasions only | 0.85 | 0.35,2.07 | 0.455 | 1.00 | 0.41,2.49 | 0.796 |
|  | Never | 2.15 | 0.81,5.74 | 0.437 | 1.37 | 0.54,3.47 | 0.664 |
| **Severe liver disease** | Daily or almost daily | 0.86 | 0.48,1.56 |  | 1.32 | 0.94,1.86 |  |
|  | Three or four times a week | 1.22 | 0.68,2.20 | 0.407 | 1.18 | 0.89,1.58 | 0.893 |
|  | Once or twice a week | 0.85 | 0.43,1.66 | 0.971 | 1.24 | 0.99,1.55 | 0.615 |
|  | One to three times a month | 4.57 | 1.94,10.76 | 0.002 | 0.77 | 0.53,1.10 | 0.246 |
|  | Special occasions only | 3.04 | 1.64,5.63 | 0.004 | 1.61 | 1.19,2.17 | 0.187 |
|  | Never | 1.52 | 0.66,3.49 | 0.260 | 1.26 | 0.86,1.84 | 0.830 |
| **NAFLD** | Daily or almost daily | 1.22 | 0.86,1.71 |  | 1.51 | 0.85,2.69 |  |
|  | Three or four times a week | 1.16 | 0.83,1.64 | 0.868 | 2.37 | 1.30,4.32 | 0.212 |
|  | Once or twice a week | 1.36 | 1.01,1.84 | 0.652 | 1.32 | 0.72,2.43 | 0.936 |
|  | One to three times a month | 1.39 | 0.89,2.16 | 0.657 | 3.15 | 1.54,6.43 | 0.007 |
|  | Special occasions only | 1.54 | 1.08,2.19 | 0.377 | 3.65 | 2.16,6.16 | 0.001 |
|  | Never | 1.29 | 0.84,1.99 | 0.788 | 1.62 | 0.75,3.50 | 0.446 |
| **Gallbladder disease** | Daily or almost daily | 1.29 | 0.92,1.80 |  | 2.83 | 1.98,4.05 |  |
|  | Three or four times a week | 0.99 | 0.73,1.33 | 0.274 | 2.60 | 1.83,3.68 | 0.521 |
|  | Once or twice a week | 1.49 | 1.17,1.90 | 0.439 | 3.32 | 2.45,4.51 | 0.666 |
|  | One to three times a month | 1.09 | 0.74,1.61 | 0.568 | 2.18 | 1.42,3.33 | 0.610 |
|  | Special occasions only | 1.55 | 1.14,2.11 | 0.383 | 3.66 | 2.43,5.52 | 0.599 |
|  | Never | 1.14 | 0.76,1.69 | 0.662 | 2.33 | 1.46,3.71 | 0.516 |
| **Pancreatic disease** | Daily or almost daily | 1.31 | 0.79,2.17 |  | 2.66 | 1.49,4.74 |  |
|  | Three or four times a week | 0.82 | 0.49,1.36 | 0.235 | 1.38 | 0.81,2.33 | 0.461 |
|  | Once or twice a week | 1.53 | 1.02,2.28 | 0.539 | 2.30 | 1.52,3.49 | 0.506 |
|  | One to three times a month | 1.56 | 0.88,2.75 | 0.572 | 1.76 | 1.01,3.07 | 0.629 |
|  | Special occasions only | 1.99 | 1.24,3.20 | 0.191 | 2.68 | 1.64,4.36 | 0.211 |
|  | Never | 1.82 | 1.05,3.14 | 0.333 | 2.04 | 1.19,3.49 | 0.416 |
|  | **Hypertension** |  |  |  |  |  |  |
| **FGID** | No | 1.44 | 1.27,1.63 | 0.594 | 1.59 | 1.40,1.79 | 0.397 |
|  | Yes | 1.51 | 1.33,1.71 |  | 1.67 | 1.48,1.88 |  |
| **PUD** | No | 1.57 | 1.19,2.06 | 0.042 | 1.30 | 1.01,1.67 | 0.001 |
|  | Yes | 1.02 | 0.74,1.39 |  | 0.76 | 0.56,1.01 |  |
| **GERD** | No | 1.49 | 1.34,1.66 | 0.514 | 1.54 | 1.39,1.71 | <0.001 |
|  | Yes | 1.41 | 1.24,1.60 |  | 1.22 | 1.09,1.37 |  |
| **IBD** | No | 0.94 | 0.59,1.50 | 0.019 | 0.92 | 0.59,1.43 | 0.050 |
|  | Yes | 2.00 | 1.31,3.06 |  | 1.58 | 1.08,2.32 |  |
| **Severe liver disease** | No | 1.21 | 0.77,1.91 | 0.306 | 1.28 | 1.08,1.52 | 0.338 |
|  | Yes | 1.61 | 1.16,2.24 |  | 1.21 | 1.01,1.44 |  |
| **NAFLD** | No | 1.30 | 1.03,1.63 | 0.749 | 2.31 | 1.47,3.62 | 0.871 |
|  | Yes | 1.35 | 1.12,1.64 |  | 2.01 | 1.49,2.70 |  |
| **Gallbladder disease** | No | 1.33 | 1.11,1.59 | 0.541 | 3.29 | 2.55,4.24 | 0.122 |
|  | Yes | 1.23 | 1.02,1.48 |  | 2.69 | 2.21,3.28 |  |
| **Pancreatic disease** | No | 1.27 | 0.95,1.72 | 0.299 | 2.34 | 1.67,3.27 | 0.395 |
|  | Yes | 1.57 | 1.20,2.05 |  | 1.98 | 1.52,2.60 |  |
|  | **Heart Failure** |  |  |  |  |  |  |
| **FGID** | No | 1.45 | 1.32,1.58 | 0.112 | 1.63 | 1.49,1.78 | 0.370 |
|  | Yes | 2.01 | 1.37,2.95 |  | 1.67 | 1.22,2.28 |  |
| **PUD** | No | 1.29 | 1.05,1.59 | 0.447 | 1.06 | 0.87,1.28 | 0.058 |
|  | Yes | 0.84 | 0.28,2.56 |  | 0.40 | 0.15,1.07 |  |
| **GERD** | No | 1.45 | 1.33,1.57 | 0.276 | 1.39 | 1.28,1.50 | 0.219 |
|  | Yes | 1.92 | 1.19,3.09 |  | 1.23 | 0.83,1.81 |  |
| **IBD** | No | 1.42 | 1.03,1.94 | 0.603 | 1.29 | 0.96,1.74 | 0.440 |
|  | Yes | 0.94 | 0.19,4.71 |  | 0.70 | 0.19,2.57 |  |
| **Severe liver disease** | No | 1.31 | 0.98,1.76 | 0.151 | 1.26 | 1.11,1.44 | 0.064 |
|  | Yes | 2.24 | 1.17,4.28 |  | 0.90 | 0.49,1.65 |  |
| **NAFLD** | No | 1.25 | 1.07,1.46 | 0.009 | 2.02 | 1.52,2.67 | 0.751 |
|  | Yes | 2.64 | 1.56,4.48 |  | 2.48 | 1.48,4.14 |  |
| **Gallbladder disease** | No | 1.31 | 1.15,1.49 | 0.160 | 2.77 | 2.35,3.27 | 0.219 |
|  | Yes | 0.81 | 0.42,1.54 |  | 4.13 | 2.57,6.63 |  |
| **Pancreatic disease** | No | 1.43 | 1.16,1.75 | 0.703 | 2.12 | 1.70,2.64 | 0.452 |
|  | Yes | 1.22 | 0.55,2.71 |  | 2.17 | 1.07,4.40 |  |
|  | **Renal Failure** |  |  |  |  |  |  |
| **FGID** | No | 1.46 | 1.33,1.60 | 0.405 | 1.63 | 1.49,1.78 | 0.135 |
|  | Yes | 1.67 | 1.24,2.25 |  | 1.62 | 1.24,2.11 |  |
| **PUD** | No | 1.28 | 1.03,1.59 | 0.858 | 1.04 | 0.85,1.27 | 0.343 |
|  | Yes | 1.21 | 0.61,2.40 |  | 0.68 | 0.37,1.24 |  |
| **GERD** | No | 1.47 | 1.36,1.60 | 0.211 | 1.42 | 1.31,1.54 | <0.001 |
|  | Yes | 1.15 | 0.80,1.66 |  | 0.91 | 0.64,1.30 |  |
| **IBD** | No | 1.39 | 1.00,1.93 | 0.965 | 1.26 | 0.93,1.71 | 0.945 |
|  | Yes | 1.42 | 0.54,3.71 |  | 1.14 | 0.46,2.85 |  |
| **Severe liver disease** | No | 1.45 | 1.09,1.92 | 0.858 | 1.28 | 1.13,1.46 | 0.043 |
|  | Yes | 1.57 | 0.68,3.62 |  | 0.90 | 0.57,1.42 |  |
| **NAFLD** | No | 1.31 | 1.12,1.53 | 0.539 | 2.31 | 1.76,3.02 | 0.068 |
|  | Yes | 1.54 | 0.97,2.44 |  | 1.23 | 0.62,2.43 |  |
| **Gallbladder disease** | No | 1.31 | 1.14,1.49 | 0.354 | 2.99 | 2.53,3.53 | 0.389 |
|  | Yes | 1.03 | 0.64,1.67 |  | 2.36 | 1.52,3.67 |  |
| **Pancreatic disease** | No | 1.38 | 1.12,1.71 | 0.384 | 2.14 | 1.71,2.68 | 0.455 |
|  | Yes | 1.82 | 1.00,3.32 |  | 2.02 | 1.11,3.68 |  |
|  | **Asthma** |  |  |  |  |  |  |
| **FGID** | No | 1.48 | 1.34,1.63 | 0.798 | 1.68 | 1.53,1.85 | 0.351 |
|  | Yes | 1.43 | 1.17,1.76 |  | 1.39 | 1.15,1.67 |  |
| **PUD** | No | 1.25 | 1.00,1.56 | 0.627 | 1.03 | 0.83,1.28 | 0.867 |
|  | Yes | 1.41 | 0.87,2.29 |  | 0.81 | 0.54,1.21 |  |
| **GERD** | No | 1.45 | 1.32,1.58 | 0.684 | 1.41 | 1.29,1.54 | 0.035 |
|  | Yes | 1.51 | 1.25,1.83 |  | 1.23 | 1.04,1.47 |  |
| **IBD** | No | 1.41 | 1.00,1.98 | 0.882 | 1.28 | 0.93,1.76 | 0.829 |
|  | Yes | 1.32 | 0.63,2.79 |  | 1.09 | 0.53,2.23 |  |
| **Severe liver disease** | No | 1.56 | 1.16,2.08 | 0.286 | 1.23 | 1.07,1.42 | 0.785 |
|  | Yes | 1.05 | 0.55,2.03 |  | 1.28 | 0.96,1.70 |  |
| **NAFLD** | No | 1.25 | 1.05,1.48 | 0.108 | 2.44 | 1.86,3.21 | 0.034 |
|  | Yes | 1.66 | 1.23,2.24 |  | 1.08 | 0.59,1.99 |  |
| **Gallbladder disease** | No | 1.25 | 1.09,1.44 | 0.389 | 2.85 | 2.38,3.42 | 0.867 |
|  | Yes | 1.45 | 1.07,1.96 |  | 3.04 | 2.23,4.14 |  |
| **Pancreatic disease** | No | 1.42 | 1.14,1.76 | 0.900 | 2.27 | 1.80,2.88 | 0.163 |
|  | Yes | 1.47 | 0.91,2.38 |  | 1.59 | 0.99,2.54 |  |
|  | **Dementia** |  |  |  |  |  |  |
| **FGID** | No | 1.45 | 1.32,1.59 | 0.177 | 1.63 | 1.49,1.77 | 0.070 |
|  | Yes | 2.13 | 1.21,3.73 |  | 1.36 | 0.94,1.99 |  |
| **PUD** | No | 1.29 | 1.05,1.58 | NA | 1.01 | 0.83,1.22 | NA |
|  | Yes | NA | NA |  | NA | NA |  |
| **GERD** | No | 1.46 | 1.34,1.58 | 0.652 | 1.37 | 1.27,1.48 | 0.645 |
|  | Yes | 1.13 | 0.52,2.43 |  | 1.54 | 0.82,2.91 |  |
| **IBD** | No | 1.40 | 1.03,1.91 | NA | 1.26 | 0.94,1.68 | NA |
|  | Yes | NA | NA |  | NA | NA |  |
| **Severe liver disease** | No | 1.44 | 1.10,1.88 | 0.402 | 1.24 | 1.10,1.41 | 0.116 |
|  | Yes | 3.78 | 0.33,43.05 |  | 0.86 | 0.33,2.21 |  |
| **NAFLD** | No | 1.34 | 1.16,1.56 | 0.357 | 2.15 | 1.67,2.77 | 0.516 |
|  | Yes | 0.78 | 0.19,3.31 |  | 1.09 | 0.32,3.67 |  |
| **Gallbladder disease** | No | 1.29 | 1.13,1.47 | 0.262 | 2.98 | 2.54,3.49 | 0.065 |
|  | Yes | 0.62 | 0.22,1.75 |  | 0.87 | 0.27,2.81 |  |
| **Pancreatic disease** | No | 1.44 | 1.18,1.76 | 0.211 | 2.19 | 1.77,2.70 | 0.137 |
|  | Yes | 0.52 | 0.11,2.45 |  | 0.74 | 0.16,3.43 |  |
|  | **MI** |  |  |  |  |  |  |
| **FGID** | No | 1.45 | 1.33,1.59 | 0.335 | 1.63 | 1.49,1.78 | 0.462 |
|  | Yes | 1.71 | 1.25,2.34 |  | 1.56 | 1.19,2.06 |  |
| **PUD** | No | 1.29 | 1.04,1.59 | 0.708 | 1.03 | 0.84,1.25 | 0.359 |
|  | Yes | 1.12 | 0.57,2.22 |  | 0.72 | 0.38,1.35 |  |
| **GERD** | No | 1.42 | 1.31,1.55 | 0.023 | 1.37 | 1.26,1.48 | 0.633 |
|  | Yes | 2.12 | 1.52,2.96 |  | 1.49 | 1.11,1.98 |  |
| **IBD** | No | 1.24 | 0.89,1.73 | 0.032 | 1.15 | 0.84,1.58 | 0.122 |
|  | Yes | 3.73 | 1.46,9.55 |  | 2.08 | 1.02,4.24 |  |
| **Severe liver disease** | No | 1.29 | 0.96,1.72 | 0.018 | 1.25 | 1.10,1.42 | 0.325 |
|  | Yes | 3.25 | 1.61,6.59 |  | 1.12 | 0.70,1.79 |  |
| **NAFLD** | No | 1.28 | 1.10,1.50 | 0.090 | 1.99 | 1.50,2.64 | 0.117 |
|  | Yes | 1.98 | 1.24,3.17 |  | 2.66 | 1.57,4.51 |  |
| **Gallbladder disease** | No | 1.28 | 1.12,1.46 | 0.948 | 2.93 | 2.47,3.47 | 0.584 |
|  | Yes | 1.30 | 0.80,2.11 |  | 2.66 | 1.76,4.02 |  |
| **Pancreatic disease** | No | 1.50 | 1.22,1.85 | 0.076 | 2.20 | 1.77,2.73 | 0.164 |
|  | Yes | 0.75 | 0.36,1.57 |  | 1.45 | 0.65,3.21 |  |
|  | **Stroke** |  |  |  |  |  |  |
| **FGID** | No | 1.45 | 1.32,1.59 | 0.146 | 1.61 | 1.48,1.76 | 0.552 |
|  | Yes | 1.92 | 1.31,2.80 |  | 1.69 | 1.20,2.38 |  |
| **PUD** | No | 1.29 | 1.05,1.59 | 0.227 | 1.04 | 0.86,1.26 | 0.025 |
|  | Yes | 0.34 | 0.04,2.65 |  | 0.12 | 0.02,0.93 |  |
| **GERD** | No | 1.47 | 1.35,1.59 | 0.372 | 1.39 | 1.29,1.51 | 0.078 |
|  | Yes | 1.18 | 0.73,1.90 |  | 0.97 | 0.63,1.50 |  |
| **IBD** | No | 1.37 | 1.00,1.88 | 0.648 | 1.21 | 0.90,1.62 | 0.897 |
|  | Yes | 1.89 | 0.46,7.78 |  | 1.95 | 0.51,7.51 |  |
| **Severe liver disease** | No | 1.43 | 1.08,1.88 | 0.540 | 1.25 | 1.10,1.42 | 0.948 |
|  | Yes | 2.04 | 0.71,5.87 |  | 1.12 | 0.65,1.93 |  |
| **NAFLD** | No | 1.32 | 1.14,1.54 | 0.670 | 2.14 | 1.65,2.78 | 0.829 |
|  | Yes | 1.58 | 0.80,3.11 |  | 1.65 | 0.73,3.74 |  |
| **Gallbladder disease** | No | 1.27 | 1.12,1.45 | 0.574 | 2.99 | 2.54,3.51 | 0.879 |
|  | Yes | 1.59 | 0.84,2.99 |  | 1.95 | 1.08,3.49 |  |
| **Pancreatic disease** | No | 1.40 | 1.14,1.71 | 0.306 | 2.12 | 1.70,2.63 | 0.595 |
|  | Yes | 2.32 | 0.93,5.77 |  | 2.15 | 0.97,4.79 |  |
|  | **COPD** |  |  |  |  |  |  |
| **FGID** | No | 1.44 | 1.31,1.58 | 0.344 | 1.64 | 1.50,1.79 | 0.585 |
|  | Yes | 1.67 | 1.25,2.23 |  | 1.57 | 1.22,2.02 |  |
| **PUD** | No | 1.28 | 1.03,1.58 | 0.711 | 1.03 | 0.84,1.26 | 0.167 |
|  | Yes | 1.10 | 0.53,2.28 |  | 0.67 | 0.35,1.27 |  |
| **GERD** | No | 1.44 | 1.32,1.56 | 0.385 | 1.41 | 1.30,1.52 | 0.033 |
|  | Yes | 1.67 | 1.20,2.33 |  | 1.14 | 0.85,1.52 |  |
| **IBD** | No | 1.26 | 0.91,1.75 | 0.064 | 1.15 | 0.85,1.57 | 0.321 |
|  | Yes | 3.30 | 1.22,8.93 |  | 2.09 | 0.92,4.72 |  |
| **Severe liver disease** | No | 1.37 | 1.02,1.83 | 0.452 | 1.23 | 1.08,1.41 | 0.543 |
|  | Yes | 1.81 | 0.93,3.51 |  | 1.29 | 0.85,1.95 |  |
| **NAFLD** | No | 1.26 | 1.08,1.48 | 0.172 | 2.19 | 1.67,2.88 | 0.708 |
|  | Yes | 1.72 | 1.14,2.61 |  | 1.70 | 0.95,3.04 |  |
| **Gallbladder disease** | No | 1.27 | 1.11,1.45 | 0.929 | 2.82 | 2.39,3.34 | 0.930 |
|  | Yes | 1.29 | 0.83,2.02 |  | 3.12 | 2.05,4.73 |  |
| **Pancreatic disease** | No | 1.48 | 1.20,1.82 | 0.157 | 2.24 | 1.79,2.80 | 0.038 |
|  | Yes | 0.85 | 0.42,1.75 |  | 1.29 | 0.65,2.57 |  |
|  | **Diabetes** |  |  |  |  |  |  |
| **FGID** | No | 1.43 | 1.30,1.57 | 0.166 | 1.57 | 1.43,1.72 | 0.175 |
|  | Yes | 1.70 | 1.36,2.12 |  | 1.88 | 1.54,2.30 |  |
| **PUD** | No | 1.38 | 1.11,1.71 | 0.057 | 1.10 | 0.90,1.35 | 0.008 |
|  | Yes | 0.73 | 0.39,1.38 |  | 0.49 | 0.27,0.87 |  |
| **GERD** | No | 1.46 | 1.34,1.59 | 0.852 | 1.40 | 1.29,1.52 | 0.016 |
|  | Yes | 1.44 | 1.12,1.86 |  | 1.17 | 0.93,1.46 |  |
| **IBD** | No | 1.39 | 0.99,1.96 | 0.983 | 1.26 | 0.91,1.75 | 0.925 |
|  | Yes | 1.41 | 0.66,3.03 |  | 1.14 | 0.60,2.17 |  |
| **Severe liver disease** | No | 1.13 | 0.80,1.59 | 0.016 | 1.17 | 1.02,1.34 | 0.374 |
|  | Yes | 2.26 | 1.46,3.49 |  | 1.56 | 1.17,2.09 |  |
| **NAFLD** | No | 1.27 | 1.07,1.51 | 0.402 | 1.90 | 1.36,2.67 | 0.207 |
|  | Yes | 1.48 | 1.11,1.96 |  | 2.26 | 1.58,3.23 |  |
| **Gallbladder disease** | No | 1.21 | 1.05,1.40 | 0.062 | 3.16 | 2.61,3.83 | 0.097 |
|  | Yes | 1.67 | 1.22,2.27 |  | 2.29 | 1.75,2.99 |  |
| **Pancreatic disease** | No | 1.36 | 1.09,1.69 | 0.335 | 2.23 | 1.76,2.84 | 0.176 |
|  | Yes | 1.74 | 1.09,2.76 |  | 1.68 | 1.09,2.59 |  |
|  | **BMI** |  |  |  |  |  |  |
| **FGID** | <30 | 1.45 | 1.30,1.62 | 0.751 | 1.51 | 1.36,1.67 | 0.100 |
|  | >=30 | 1.49 | 1.28,1.74 |  | 1.87 | 1.62,2.17 |  |
| **PUD** | <30 | 1.47 | 1.15,1.89 | 0.038 | 1.09 | 0.87,1.36 | 0.155 |
|  | >=30 | 0.92 | 0.64,1.33 |  | 0.78 | 0.55,1.12 |  |
| **GERD** | <30 | 1.45 | 1.32,1.60 | 0.984 | 1.35 | 1.23,1.48 | 0.784 |
|  | >=30 | 1.46 | 1.26,1.69 |  | 1.42 | 1.24,1.62 |  |
| **IBD** | <30 | 1.38 | 0.96,1.98 | 0.855 | 1.38 | 0.97,1.95 | 0.630 |
|  | >=30 | 1.48 | 0.80,2.76 |  | 0.97 | 0.58,1.65 |  |
| **Severe liver disease** | <30 | 1.39 | 0.98,1.98 | 0.737 | 1.15 | 0.98,1.36 | 0.187 |
|  | >=30 | 1.53 | 1.02,2.30 |  | 1.34 | 1.10,1.62 |  |
| **NAFLD** | <30 | 1.26 | 1.02,1.56 | 0.663 | 2.21 | 1.59,3.08 | 0.468 |
|  | >=30 | 1.35 | 1.10,1.66 |  | 1.89 | 1.30,2.74 |  |
| **Gallbladder disease** | <30 | 1.20 | 1.02,1.43 | 0.374 | 2.88 | 2.31,3.60 | 0.914 |
|  | >=30 | 1.36 | 1.11,1.65 |  | 2.77 | 2.24,3.43 |  |
| **Pancreatic disease** | <30 | 1.19 | 0.92,1.54 | 0.019 | 1.91 | 1.45,2.51 | 0.116 |
|  | >=30 | 1.94 | 1.41,2.67 |  | 2.45 | 1.77,3.39 |  |
|  | **Townsend Deprivation Index** |  |  |  |  |  |  |
| **FGID** | <mean | 1.44 | 1.28,1.63 | 0.648 | 1.68 | 1.49,1.88 | 0.790 |
|  | >=mean | 1.50 | 1.32,1.72 |  | 1.54 | 1.36,1.75 |  |
| **PUD** | <mean | 1.12 | 0.84,1.50 | 0.233 | 0.89 | 0.67,1.17 | 0.509 |
|  | >=mean | 1.44 | 1.08,1.92 |  | 1.05 | 0.81,1.36 |  |
| **GERD** | <mean | 1.40 | 1.25,1.57 | 0.432 | 1.27 | 1.15,1.42 | 0.105 |
|  | >=mean | 1.50 | 1.33,1.69 |  | 1.48 | 1.32,1.66 |  |
| **IBD** | <mean | 1.56 | 1.02,2.36 | 0.449 | 1.21 | 0.82,1.78 | 0.874 |
|  | >=mean | 1.22 | 0.77,1.93 |  | 1.27 | 0.82,1.96 |  |
| **Severe liver disease** | <mean | 1.38 | 0.93,2.05 | 0.759 | 1.10 | 0.93,1.31 | 0.058 |
|  | >=mean | 1.50 | 1.04,2.15 |  | 1.39 | 1.16,1.66 |  |
| **NAFLD** | <mean | 1.18 | 0.95,1.46 | 0.176 | 2.40 | 1.65,3.50 | 0.775 |
|  | >=mean | 1.45 | 1.18,1.77 |  | 1.84 | 1.32,2.56 |  |
| **Gallbladder disease** | <mean | 1.21 | 1.01,1.45 | 0.449 | 2.89 | 2.30,3.62 | 0.570 |
|  | >=mean | 1.34 | 1.11,1.62 |  | 2.77 | 2.25,3.43 |  |
| **Pancreatic disease** | <mean | 1.13 | 0.84,1.52 | 0.036 | 1.77 | 1.31,2.41 | 0.045 |
|  | >=mean | 1.74 | 1.32,2.29 |  | 2.43 | 1.82,3.24 |  |

HR: hazard ratio; CI: confidence interval; Pi: P value for interaction.

Weighted HRs after IPTW and 95% CIs are presented.

Supplementary Table 11. Hazard ratio of digestive outcomes in COVID-19 group, the contemporary and historical control by sex

| **Outcome** | **Subgroup** | **COVID-19 vs Contemporary control** | | | **COVID-19 vs Historical control** | | |
| --- | --- | --- | --- | --- | --- | --- | --- |
|  |  | **HR** | **95% CI** | **Pi** | **HR** | **95% CI** | **Pi** |
| **FGID** | Male | 1.61 | 1.41,1.83 | 0.074 | 1.86 | 1.64,2.10 | 0.068 |
|  | Female | 1.37 | 1.21,1.54 |  | 1.44 | 1.28,1.62 |  |
| **PUD** | Male | 1.32 | 1.00,1.74 | 0.667 | 1.03 | 0.80,1.33 | 0.595 |
|  | Female | 1.20 | 0.89,1.63 |  | 0.93 | 0.70,1.23 |  |
| **GERD** | Male | 1.46 | 1.29,1.65 | 0.971 | 1.45 | 1.29,1.63 | 0.367 |
|  | Female | 1.46 | 1.31,1.63 |  | 1.32 | 1.19,1.47 |  |
| **IBD** | Male | 1.65 | 1.08,2.52 | 0.286 | 1.36 | 0.93,1.99 | 0.377 |
|  | Female | 1.17 | 0.74,1.85 |  | 1.12 | 0.71,1.75 |  |
| **Severe liver disease** | Male | 1.61 | 1.16,2.22 | 0.332 | 1.37 | 1.13,1.65 | 0.371 |
|  | Female | 1.20 | 0.75,1.92 |  | 1.16 | 0.99,1.37 |  |
| **NAFLD** | Male | 1.21 | 0.98,1.49 | 0.227 | 2.32 | 1.70,3.16 | 0.659 |
|  | Female | 1.47 | 1.19,1.81 |  | 1.69 | 1.12,2.55 |  |
| **Gallbladder disease** | Male | 1.32 | 1.09,1.61 | 0.723 | 2.92 | 2.35,3.62 | 0.580 |
|  | Female | 1.26 | 1.06,1.49 |  | 2.88 | 2.30,3.62 |  |
| **Pancreatic disease** | Male | 1.17 | 0.88,1.57 | 0.069 | 1.92 | 1.40,2.61 | 0.276 |
|  | Female | 1.69 | 1.29,2.22 |  | 2.32 | 1.74,3.08 |  |

Supplementary Table 12. Baseline characteristics of COVID-19 group and historical controls before weighting

| Characteristics | COVID-19 group  (n = 112 311） | Historical controls  (n = 370 979) | SMD |
| --- | --- | --- | --- |
| Age, mean(SD), years | 54.43 (8.19) | 56.94 (7.97) | 0.312 |
| Sex, female, n(%) | 50 816 (45.2) | 166 904 (45.0) | 0.005 |
| Ethnicity, White, n(%) | 106 315 (94.7) | 350 562 (94.5) | 0.007 |
| Household income |  |  | 0.201 |
| <18 000, n(%) | 19 465 (17.3) | 87 290 (23.5) |  |
| 18 000-30 999, n(%) | 26 102 (23.2) | 97 269 (26.2) |  |
| 31 000-51 999, n(%) | 31 853 (28.4) | 94 903 (25.6) |  |
| 52 000-100 000, n(%) | 27 297 (24.3) | 71 710 (19.3) |  |
| >100 000, n(%) | 7 594 ( 6.8) | 19 807 ( 5.3) |  |
| Deprivation index, mean(SD) | -1.38 (3.00) | -1.30 (3.10) | 0.028 |
| BMI, mean(SD), kg/m^2^ | 27.47 (4.82) | 27.39 (4.76) | 0.018 |
| Alcohol consumption |  |  | 0.104 |
| Daily or almost daily, n(%) | 21 925 (19.5) | 75 808 (20.4) |  |
| Three or four times a week, n(%) | 28 027 (25.0) | 84 054 (22.7) |  |
| Once or twice a week, n(%) | 30 734 (27.4) | 94 503 (25.5) |  |
| One to three times a month, n(%) | 12 790 (11.4) | 41 435 (11.2) |  |
| Special occasions only or never, n(%) | 11 298 (10.1) | 44 263 (11.9) |  |
| Never, n(%) | 7 537 ( 6.7) | 30 916 ( 8.3) |  |
| Smoking status |  |  | 0.062 |
| Never smoker, n(%) | 63 505 (56.5) | 204 121 (55.0) |  |
| Previous smoker, n(%) | 38 875 (34.6) | 127 264 (34.3) |  |
| Current smoker, n(%) | 9 931 ( 8.8) | 39 594 (10.7) |  |
| Physical activity, mean(SD), MET minutes/week | 2536.46 (2603.69) | 2683.22 (2740.86) | 0.055 |
| Comorbidities |  |  |  |
| Hypertension, n(%) | 38 023 (33.9) | 139 121 (37.5) | 0.076 |
| Diabetes, n(%) | 8 384 ( 7.5) | 30 043 ( 8.1) | 0.024 |
| Renal failure, n(%) | 4 646 ( 4.1) | 17 084 ( 4.6) | 0.023 |
| Myocardial infarction, n(%) | 4 557 ( 4.1) | 17 795 ( 4.8) | 0.036 |
| Stroke, n(%) | 2 706 ( 2.4) | 11 228 ( 3.0) | 0.038 |
| COPD, n(%) | 4 377 ( 3.9) | 17 197 ( 4.6) | 0.037 |
| Asthma, n(%) | 17 231 (15.3) | 50 073 (13.5) | 0.053 |
| Heart failure, n(%) | 2 356 ( 2.1) | 10 127 ( 2.7) | 0.041 |
| Dementia, n(%) | 1 060 ( 0.9) | 3 788 ( 1.0) | 0.008 |
| History of previous digestive diseases, n(%) | 37 375 (33.3) | 109 941 (29.6) | 0.079 |

SMD: standard mean difference; BMI: body mass index; MET: metabolic equivalent of task; COPD: chronic obstructive pulmonary disease; SD: standard deviation

Supplementary Table 13. Baseline characteristics of COVID-19 group and historical controls after weighting

| Characteristics | COVID-19 group  (n = 112 311) | Historical controls  (n = 370 979) | SMD |
| --- | --- | --- | --- |
| Age, mean(SD), years | 56.2(8.1) | 56.4(8.1) | <0.001 |
| Sex, female, n(%) | 61 546(54.8) | 203 667(54.9) | 0.008 |
| Ethnicity, White, n(%) | 10 6134(94.5) | 350 575(94.5) | 0.001 |
| Household income |  |  | 0.003 |
| <18 000, n(%) | 24 371(21.7) | 81 986(22.1) |  |
| 18 000-30 999, n(%) | 28 527(25.4) | 94 600(25.5) |  |
| 31 000-51 999, n(%) | 29 650(26.4) | 97 196(26.2) |  |
| 52 000-100 000, n(%) | 23 361(20.8) | 76 051(20.5) |  |
| >100 000, n(%) | 6 514(5.8) | 21 146(5.7) |  |
| Deprivation index, mean(SD) | -1.3(3.0) | -1.3(3.1) | 0.001 |
| BMI, mean(SD), kg/m^2^ | 27.4(4.7) | 27.4(4.8) | 0.001 |
| Alcohol consumption |  |  | 0.005 |
| Daily or almost daily, n(%) | 22 799(20.3) | 74 938(20.2) |  |
| Three or four times a week, n(%) | 26 168(23.3) | 86 067(23.2) |  |
| Once or twice a week, n(%) | 29 089(25.9) | 96 084(25.9) |  |
| One to three times a month, n(%) | 12 579(11.2) | 415 50(11.2) |  |
| Special occasions only or never, n(%) | 12 803(11.4) | 42 663(11.5) |  |
| Never, n(%) | 8 873(7.9) | 29 678(8) |  |
| Smoking status |  |  | 0.001 |
| Never smoker, n(%) | 62 557(55.7) | 205 522(55.4) |  |
| Previous smoker, n(%) | 38 523(34.3) | 127 617(34.4) |  |
| Current smoker, n(%) | 11 343(10.1) | 37 840(10.2) |  |
| Physical activity, mean(SD), MET minutes/week | 2645.2(2702.9) | 2648.5(2711.0) | 0.002 |
| Comorbidities |  |  |  |
| Hypertension, n(%) | 40 657(36.2) | 136 149(36.7) | 0.001 |
| Diabetes, n(%) | 8 648(7.7) | 29 678(8) | 0.001 |
| Renal failure, n(%) | 4 717(4.2) | 16 694(4.5) | 0.001 |
| Myocardial infarction, n(%) | 4 829(4.3) | 17 065(4.6) | 0.001 |
| Stroke, n(%) | 3 032(2.7) | 10758(2.9) | 0.003 |
| COPD, n(%) | 5 054(4.5) | 16 694(4.5) | 0.004 |
| Asthma, n(%) | 15 611(13.9) | 51 566(13.9) | 0.001 |
| Heart failure, n(%) | 2 583(2.3) | 9 645(2.6) | 0.011 |
| Dementia, n(%) | 786(0.7) | 3 710(1) | 0.003 |
| History of previous digestive diseases, n(%) | 36 950(32.9) | 113 149(30.5) | 0.005 |

SMD: standard mean difference; BMI: body mass index; MET: metabolic equivalent of task; COPD: chronic obstructive pulmonary disease; SD: standard deviation

Supplementary Table 14. Baseline characteristics of COVID-19 group and historical controls by severity of COVID-19 before weighting

| Characteristics | Non-hospitalized COVID  (n= 104 201) | Hospitalized COVID  (n= 7 523) | Severe COVID  (n= 588) | Historical controls  (n = 370 979) | SMD | | |
| --- | --- | --- | --- | --- | --- | --- | --- |
|  |  |  |  |  | Non-hospitalized COVID and historical controls | Hospitalized COVID and historical controls | Severe COVID and historical controls |
| Age, mean(SD), years | 54.02(8.09) | 59.84(7.55) | 57.78(7.99) | 56.94(7.97) | 0.365 | 0.374 | 0.105 |
| Sex, female, n(%) | 57 959(55.6) | 3 324(44.2) | 212(36.1) | 166 904 (45.0) | 0.012 | 0.218 | 0.388 |
| Ethnicity, White, n(%) | 98 873(94.9) | 6 951(92.4) | 492(83.7) | 350 562 (94.5) |  |  |  |
| Household income |  |  |  |  | 0.244 | 0.322 | 0.378 |
| <18 000, n(%) | 16 497(15.8) | 2 741(36.4) | 227(38.6) | 87 290 (23.5) |  |  |  |
| 18 000-30 999, n(%) | 23 945(23.0) | 2 003(26.6) | 154(26.2) | 97 269 (26.2) |  |  |  |
| 31 000-51 999, n(%) | 30 165(28.9) | 1 567(20.8) | 122(20.7) | 94 903 (25.6) |  |  |  |
| 52 000-100 000, n(%) | 26 266(25.2) | 961(12.8) | 70(11.9) | 71 710 (19.3) |  |  |  |
| >100 000, n(%) | 7 328(7.0) | 251(3.3) | 15(2.6) | 19 807 ( 5.3) |  |  |  |
| Deprivation index, mean(SD) | -1.46(2.95) | -0.43(3.40) | 0.16(3.54) | -1.30 (3.10) | 0.054 | 0.268 | 0.437 |
| BMI, mean(SD), kg/m^2^ | 27.33(4.72) | 29.23(5.63) | 30.61(5.78) | 27.39 (4.76) | 0.012 | 0.353 | 0.608 |
| Alcohol consumption |  |  |  |  | 0.129 | 0.204 | 0.312 |
| Daily or almost daily, n(%) | 20 416(19.6) | 1 414(18.8) | 96(16.3) | 75 808 (20.4) |  |  |  |
| Three or four times a week, n(%) | 26 582(25.5) | 1353(18.0) | 92(15.6) | 84 054 (22.7) |  |  |  |
| Once or twice a week, n(%) | 28 762(27.6) | 1 825(24.3) | 147(25.0) | 94 503 (25.5) |  |  |  |
| One to three times a month, n(%) | 11 927(11.4) | 799(10.6) | 64(10.9) | 41 435 (11.2) |  |  |  |
| Special occasions only or never, n(%) | 10 034(9.6) | 1 168(15.5) | 96(16.3) | 44 263 (11.9) |  |  |  |
| Never, n(%) | 6 480(6.2) | 964(12.8) | 93(15.8) | 30 916 ( 8.3) |  |  |  |
| Smoking status |  |  |  |  | 0.080 | 0.217 | 0.252 |
| Never smoker, n(%) | 59 921(57.5) | 3 335(44.3) | 250(42.5) | 204 121 (55.0) |  |  |  |
| Previous smoker, n(%) | 35 500(34.1) | 3 115(41.4) | 260(44.2) | 127 264 (34.3) |  |  |  |
| Current smoker, n(%) | 8 780(8.4) | 1 073(14.3) | 78(13.3) | 39 594 (10.7) |  |  |  |
| Physical activity, mean(SD), MET minutes/week | 2529.22  (2585.74) | 2623.26  (2813.78) | 2707.49  (2935.05) | 2683.22 (2740.86) | 0.058 | 0.022 | 0.009 |
| Comorbidities |  |  |  |  |  |  |  |
| Hypertension, n(%) | 33 161(31.8) | 4 523(60.1) | 339(57.7) | 139 121 (37.5) | 0.12 | 0.465 | 0.412 |
| Diabetes, n(%) | 6 579(6.3) | 1 678(22.3) | 127(21.6) | 30 043 ( 8.1) | 0.069 | 0.404 | 0.387 |
| Renal failure, n(%) | 3 515(3.4) | 1 050(14.0) | 81(13.8) | 17 084 ( 4.6) | 0.063 | 0.327 | 0.322 |
| Myocardial infarction, n(%) | 3 599(3.5) | 890(11.8) | 68(11.6) | 17 795 ( 4.8) | 0.068 | 0.257 | 0.249 |
| Stroke, n(%) | 2 056(2.0) | 623(8.3) | 27(4.6) | 11 228 ( 3.0) | 0.068 | 0.229 | 0.082 |
| COPD, n(%) | 3 174(3.0) | 1 123(14.9) | 80(13.6) | 17 197 ( 4.6) | 0.083 | 0.352 | 0.315 |
| Asthma, n(%) | 15 561(14.9) | 1 534(20.4) | 136(23.1) | 50 073 (13.5) | 0.041 | 0.185 | 0.251 |
| Heart failure, n(%) | 1 607(1.5) | 714(9.5) | 35(6.0) | 10 127 ( 2.7) | 0.082 | 0.285 | 0.159 |
| Dementia, n(%) | 732(0.7) | 321(4.3) | 7(1.2) | 3 788 ( 1.0) | 0.034 | 0.203 | 0.016 |
| History of previous digestive diseases, n(%) | 33 024(31.7) | 4 085(54.3) | 266(45.2) | 109 941 (29.6) | 0.045 | 0.516 | 0.327 |

SMD: standard mean difference; BMI: body mass index; MET: metabolic equivalent of task; COPD: chronic obstructive pulmonary disease; SD: standard deviation

Supplementary Table 15. Baseline characteristics of COVID-19 group and historical controls by severity of COVID-19 after weighting

| Characteristics | Non-hospitalized COVID  (n= 104 201) | Hospitalized COVID  (n= 7 523) | Severe COVID  (n= 588) | Historical controls  (n = 370 979) | SMD | | |
| --- | --- | --- | --- | --- | --- | --- | --- |
|  |  |  |  |  | Non-hospitalized COVID and historical controls | Hospitalized COVID and historical controls | Severe COVID and historical controls |
| Age, mean(SD), years | 54.6(8.2) | 58.6(7.9) | 57.7(7.6) | 56.4(8.1) | **0.276** | **0.207** | **0.103** |
| Sex, female, n(%) | 57 623(55.3) | 4 010(53.3) | 293(49.9) | 203 667(54.9) | 0.003 | 0.030 | **0.103** |
| Ethnicity, White, n(%) | 98 470(94.5) | 7 094(94.3) | 550(93.6) | 350 575(94.5) | 0.002 | 0.007 | 0.038 |
| Household income |  |  |  |  | 0.001 | 0.039 | 0.074 |
| <18 000, n(%) | 22 716(21.8) | 1 881(25) | 149(25.4) | 81 986(22.1) |  |  |  |
| 18 000-30 999, n(%) | 26 571(25.5) | 1 964(26.1) | 163(27.7) | 94 600(25.5) |  |  |  |
| 31 000-51 999, n(%) | 27 405(26.3) | 1 941(25.8) | 142(24.1) | 97 196(26.2) |  |  |  |
| 52 000-100 000, n(%) | 21 570(20.7) | 1 362(18.1) | 102(17.3) | 76 051(20.5) |  |  |  |
| >100 000, n(%) | 5 939(5.7) | 369(4.9) | 32(5.5) | 21 146(5.7) |  |  |  |
| Deprivation index, mean(SD) | -1.3(3.0) | -1.2(3.1) | -1.1(3.2) | -1.3(3.1) | 0.003 | 0.018 | 0.064 |
| BMI, mean(SD), kg/m^2^ | 27.4(4.7) | 27.6(4.9) | 28.3(4.7) | 27.4(4.8) | <0.001 | 0.038 | **0.200** |
| Alcohol consumption |  |  |  |  | 0.001 | 0.037 | **0.144** |
| Daily or almost daily, n(%) | 21 153(20.3) | 1 550(20.6) | 108(18.4) | 74 938(20.2) |  |  |  |
| Three or four times a week, n(%) | 24 279(23.3) | 1 617(21.5) | 114(19.4) | 86 067(23.2) |  |  |  |
| Once or twice a week, n(%) | 27 092(26) | 1 866(24.8) | 152(25.8) | 96 084(25.9) |  |  |  |
| One to three times a month, n(%) | 11 671(11.2) | 873(11.6) | 77(13.1) | 415 50(11.2) |  |  |  |
| Special occasions only or never, n(%) | 11 879(11.4) | 955(12.7) | 68(11.6) | 42 663(11.5) |  |  |  |
| Never, n(%) | 8 128(7.8) | 662(8.8) | 68(11.6) | 29 678(8) |  |  |  |
| Smoking status |  |  |  |  | 0.005 | 0.043 | 0.072 |
| Never smoker, n(%) | 58 040(55.7) | 3 987(53) | 325(55.2) | 205 522(55.4) |  |  |  |
| Previous smoker, n(%) | 357 41(34.3) | 2 648(35.2) | 189(32.1) | 127 617(34.4) |  |  |  |
| Current smoker, n(%) | 10 420(10) | 895(11.9) | 75(12.7) | 37 840(10.2) |  |  |  |
| Physical activity, mean(SD), MET minutes/week | 2653.2(2722.8) | 2636.5(2676.4) | 2675.2(2777.8) | 2648.5(2711.0) | 0.001 | 0.017 | 0.003 |
| Comorbidities |  |  |  |  |  |  |  |
| Hypertension, n(%) | 37 825(36.3) | 2 904(38.6) | 260(44.2) | 136 149(36.7) | <0.001 | 0.014 | **0.135** |
| Diabetes, n(%) | 8 023(7.7) | 677(9) | 65(11.1) | 29 678(8) | 0.001 | 0.023 | 0.100 |
| Renal failure, n(%) | 4 585(4.4) | 399(5.3) | 36(6.1) | 16 694(4.5) | 0.001 | 0.024 | 0.066 |
| Myocardial infarction, n(%) | 4 689(4.5) | 406(5.4) | 39(6.6) | 17 065(4.6) | <0.001 | 0.021 | 0.077 |
| Stroke, n(%) | 2 918(2.8) | 271(3.6) | 22(3.7) | 10758(2.9) | 0.002 | 0.026 | 0.038 |
| COPD, n(%) | 3 647(3.5) | 670(8.9) | 56(9.5) | 16 694(4.5) | 0.048 | **0.165** | **0.190** |
| Asthma, n(%) | 14 380(13.8) | 1 053(14) | 97(16.5) | 51 566(13.9) | <0.001 | 0.010 | 0.085 |
| Heart failure, n(%) |  |  |  | 9 645(2.6) |  |  |  |
| Dementia, n(%) | 1 042(1) | 113(1.5) | 4(0.6) | 3 710(1) | 0.005 | 0.040 | 0.051 |
| History of previous digestive diseases, n(%) | 31 573(30.3) | 23 47(31.2) | 203(34.6) | 113 149(30.5) | 0.004 | 0.022 | **0.106** |

SMD: standard mean difference; BMI: body mass index; MET: metabolic equivalent of task; COPD: chronic obstructive pulmonary disease; SD: standard deviation

Supplementary Table 16. Hazard ratio of digestive outcomes in COVID-19 group and the historical control by severity of COVID-19

| Outcome | HR (95% CI) | P value |
| --- | --- | --- |
| FGID |  |  |
| Non-hospitalized COVID | 1.14(1.03,1.26) | **0.013** |
| Hospitalized COVID | 4.00(3.24,4.95) | **<0.001** |
| Severe COVID | 2.66(1.38,5.12) | **0.003** |
| Peptic ulcer |  |  |
| Non-hospitalized COVID | 1.09(0.88,1.36) | 0.422 |
| Hospitalized COVID | 1.34(0.74,2.45) | 0.338 |
| Severe COVID | 0.70(0.23,2.11) | 0.528 |
| Gastro-oesophageal reflux disease |  |  |
| Non-hospitalized COVID | 1.29(1.18,1.40) | **<0.001** |
| Hospitalized COVID | 1.85(1.43,2.39) | **<0.001** |
| Severe COVID | 2.28(1.06,4.92) | **0.036** |
| Inflammatory bowel disease |  |  |
| Non-hospitalized COVID | 1.20(0.85,1.70) | 0.304 |
| Hospitalized COVID | 1.47(0.61,3.54) | 0.387 |
| Severe COVID | NA | NA |
| Gallbladder disease |  |  |
| Non-hospitalized COVID | 1.12(0.97,1.29) | 0.133 |
| Hospitalized COVID | 2.43(1.79,3.29) | **<0.001** |
| Severe COVID | 2.92(1.01,8.48) | **0.049** |
| Severe liver disease |  |  |
| Non-hospitalized COVID | 1.02(0.73,1.41) | 0.912 |
| Hospitalized COVID | 3.12(1.89,5.14) | **<0.001** |
| Severe COVID | 3.08(0.38,25.25) | 0.294 |
| Non-alcoholic fatty liver disease |  |  |
| Non-hospitalized COVID | 1.73(1.45,2.05) | **<0.001** |
| Hospitalized COVID | 2.49(1.63,3.81) | **<0.001** |
| Severe COVID | 2.03(0.54,7.62) | 0.297 |
| Pancreatic disease |  |  |
| Non-hospitalized COVID | 1.28(1.01,1.63) | **0.045** |
| Hospitalized COVID | 3.71(2.45,5.62) | **<0.001** |
| Severe COVID | 4.65(0.85,25.42) | 0.076 |

HR: hazard ratio; CI: confidence interval;

Outcomes were ascertained 30 days after the COVID-19-positive test until the end of follow-up. Weighted HRs after IPTW and 95% CIs are presented.

Supplementary Table 17. Baseline characteristics of COVID-19 group and histotical controls in the sensitive analysis restricting to the period before vaccination was available before weighting.

| Characteristics | COVID-19 group  (n = 8 431) | Historical controls  (n = 370 979) | SMD |
| --- | --- | --- | --- |
| Age, mean(SD), years | 54.39(8.82) | 56.94(7.97) | 0.304 |
| Sex, female, n(%) | 4 280(50.8) | 204 075(55.0) | 0.085 |
| Ethnicity, White, n(%) | 7 771(92.2) | 350 562(94.5) | 0.093 |
| Household income |  |  | 0.064 |
| <18 000, n(%) | 2 147(25.5) | 87 290(23.5) |  |
| 18 000-30 999, n(%) | 2 119(25.1) | 97 269(26.2) |  |
| 31 000-51 999, n(%) | 2 236(26.5) | 94 903(25.6) |  |
| 52 000-100 000, n(%) | 1 546(18.3) | 71 710(19.3) |  |
| >100 000, n(%) | 383(4.5) | 19 807(5.3) |  |
| Deprivation index, mean(SD) | -0.73(3.27) | -1.30(3.10) | 0.178 |
| BMI, mean(SD), kg/m^2^ | 28.43(5.07) | 27.39(4.76) | 0.211 |
| Alcohol consumption |  |  | 0.115 |
| Daily or almost daily, n(%) | 1 408(16.7) | 75 808(20.4) |  |
| Three or four times a week, n(%) | 1 843(21.9) | 84 054(22.7) |  |
| Once or twice a week, n(%) | 2 394(28.4) | 94 503(25.5) |  |
| One to three times a month, n(%) | 965(11.4) | 41 435(11.2) |  |
| Special occasions only or never, n(%) | 1 004(11.9) | 44 263(11.9) |  |
| Never, n(%) | 817(9.7) | 30 916(8.3) |  |
| Smoking status |  |  | 0.076 |
| Never smoker, n(%) | 4 322(51.3) | 204 121(55.0) |  |
| Previous smoker, n(%) | 3 115(36.9) | 127 264(34.3) |  |
| Current smoker, n(%) | 994(11.8) | 39 594(10.7) |  |
| Physical activity, mean(SD), MET minutes/week | 2734.76(2828.44) | 2683.22(2740.86) | 0.019 |
| Comorbidities |  |  |  |
| Hypertension, n(%) | 3 411(40.5) | 139 121(37.5) | 0.061 |
| Diabetes, n(%) | 1 005(11.9) | 30 043(8.1) | 0.128 |
| Renal failure, n(%) | 573(6.8) | 17 084(4.6) | 0.095 |
| Myocardial infarction, n(%) | 547(6.5) | 17 795(4.8) | 0.073 |
| Stroke, n(%) | 398(4.7) | 11 228(3.0) | 0.088 |
| COPD, n(%) | 616(7.3) | 17 197(4.6) | 0.113 |
| Asthma, n(%) | 1 382(16.4) | 50 073(13.5) | 0.081 |
| Heart failure, n(%) | 385(4.6) | 10 127(2.7) | 0.098 |
| Dementia, n(%) | 345(4.1) | 3 788(1.0) | 0.195 |
| History of previous digestive diseases, n(%) | 3 286(39.0) | 109 941(29.6) | 0.198 |

SMD: standard mean difference; BMI: body mass index; MET: metabolic equivalent of task; COPD: chronic obstructive pulmonary disease; SD: standard deviation

Supplementary Table 18. Baseline characteristics of COVID-19 group and historical controls in the sensitive analysis restricting to the period before vaccination was available after weighting.

| Characteristics | COVID-19 group  (n = 8 431) | Historical controls  (n = 370 979) | SMD |
| --- | --- | --- | --- |
| Age, mean(SD), years | 56.7(8.5) | 56.9(8.0) | 0.002 |
| Sex, female, n(%) | 4 519(53.6) | 203 667(54.9) | 0.034 |
| Ethnicity, White, n(%) | 7 934(94.1) | 350 204(94.4) | 0.007 |
| Household income |  |  | 0.031 |
| <18 000, n(%) | 2 049(24.3) | 87 551(23.6) |  |
| 18 000-30 999, n(%) | 2 167(25.7) | 97 196(26.2) |  |
| 31 000-51 999, n(%) | 2 150(25.5) | 94 971(25.6) |  |
| 52 000-100 000, n(%) | 1 602(19) | 71 599(19.3) |  |
| >100 000, n(%) | 464(5.5) | 19 662(5.3) |  |
| Deprivation index, mean(SD) | -1.2(3.1) | -1.3(3.1) | 0.034 |
| BMI, mean(SD), kg/m^2^ | 27.6(4.6) | 27.4(4.8) | 0.043 |
| Alcohol consumption |  |  | 0.020 |
| Daily or almost daily, n(%) | 1 728(20.5) | 75 680(20.4) |  |
| Three or four times a week, n(%) | 1 889(22.4) | 83 841(22.6) |  |
| Once or twice a week, n(%) | 2 125(25.2) | 94 600(25.5) |  |
| One to three times a month, n(%) | 936(11.1) | 41 550(11.2) |  |
| Special occasions only or never, n(%) | 1 012(12) | 44 147(11.9) |  |
| Never, n(%) | 742(8.8) | 31 162(8.4) |  |
| Smoking status |  |  | 0.039 |
| Never smoker, n(%) | 4 553(54) | 203 667(54.9) |  |
| Previous smoker, n(%) | 2 900(34.4) | 127 617(34.4) |  |
| Current smoker, n(%) | 978(11.6) | 39 695(10.7) |  |
| Physical activity, mean(SD), MET minutes/week | 2687.5 (2742.4) | 2684.3(2744.3) | 0.004 |
| Comorbidities |  |  |  |
| Hypertension, n(%) | 3 238(38.4) | 139 488(37.6) | 0.032 |
| Diabetes, n(%) | 700(8.3) | 30 420(8.2) | 0.022 |
| Renal failure, n(%) | 363(4.3) | 17 436(4.7) | 0.008 |
| Myocardial infarction, n(%) | 379(4.5) | 17 807(4.8) | 0.005 |
| Stroke, n(%) | 261(3.1) | 11 500(3.1) | 0.029 |
| COPD, n(%) | 582(6.9) | 17 436(4.7) | 0.095 |
| Asthma, n(%) | 1 155(13.7) | 50 453(13.6) | 0.010 |
| Heart failure, n(%) | 304(3.6) | 10 387(2.8) | 0.060 |
| Dementia, n(%) | 67(0.8) | 4 081(1.1) | 0.015 |
| History of previous digestive diseases, n(%) | 3 263(38.7) | 110 552(29.8) | 0.023 |

SMD: standard mean difference; BMI: body mass index; MET: metabolic equivalent of task; COPD: chronic obstructive pulmonary disease; SD: standard deviation
